# Genetic correlates of phenotypic heterogeneity in autism

**DOI:** 10.1101/2020.07.21.20159228

**Authors:** Varun Warrier, Xinhe Zhang, Patrick Reed, Alexandra Havdahl, Tyler M Moore, Freddy Cliquet, Claire S Leblond, Thomas Rolland, Anders Rosengren, EU-AIMS-LEAP, iPSYCH-Autism Working Group, Spectrum 10K and APEX Consortium, David H Rowitch, Matthew E Hurles, Daniel H Geschwind, Anders D Børglum, Elise B Robinson, Jakob Grove, Hilary C Martin, Thomas Bourgeron, Simon Baron-Cohen

**Author notes:** Correspondence to Varun Warrier. Group authorship: details at the end of the manuscript. Shared senior authors (Thomas Bourgeron, Simon Baron-Cohen).

## Abstract

The substantial phenotypic heterogeneity in autism limits our understanding of its genetic aetiology. To address this gap, we investigated genetic differences between autistic individuals (N_max_ = 12,893) based on core (i.e., social communication difficulties, and restricted and repetitive behaviours) and associated features of autism, co-occurring developmental disabilities (e.g. language, motor, and intellectual developmental disabilities and delays), and sex. We conducted a comprehensive factor analysis of core autism features in autistic individuals and identified six factors. Common genetic variants including autism polygenic scores (PGS) were associated with the core factors but *de novo* variants were not, even though the latent factor structure was similar between carriers and non-carriers of *de novo* variants. We identify that increasing autism PGS decrease the likelihood of co-occurring developmental disabilities in autistic individuals, which reflects both a true protective effect and additivity between rare and common variants. Furthermore in autistic individuals without co-occurring intellectual disability (ID), autism PGS are overinherited by autistic females compared to males. Finally, we observe higher SNP heritability for males and autistic individuals without ID, but found no robust differences in SNP heritability by the level of core autism features. Deeper phenotypic characterisation will be critical to determining how the complex underlying genetics shapes cognition, behaviour, and co-occurring conditions in autism.

## Introduction

The core diagnostic criteria for Autism Spectrum Disorder (henceforth, autism) consist of social communication difficulties, unusually restricted and repetitive behaviour, and sensory difficulties that are present early in life and affect social, occupational, and other important domains of functioning.^1, 2^ However, these criteria are reasonably broad such that two individuals with very different phenotypic features, co-occurring conditions, support needs or outcomes may both be diagnosed as autistic^1, 3,1#^. The advantage of broad diagnostic criteria is that they allow for different individuals to access important clinical, educational, and social support services. Nonetheless, this immense heterogeneity is a challenge for research that aims to understand the causes of autism and develop evidence-based support strategies for autistic people.

Heterogeneity in autism can arise from multiple, partly overlapping sources. This includes differences in core diagnostic features (core features)^1, 3, 4^, and associated features such as IQ, adaptive behaviour, and motor coordination, all of which have an impact on life outcomes.^3, 5, 6^ Furthermore, sex and gender^7, 8^ and co-occurring intellectual disability (ID) and developmental, behavioural, and medical conditions^9, 10^ can alter the presentation and measurement of core autism features. Whilst a few studies have attempted to investigate the genetic influences on this heterogeneity^11–18^, substantial gaps remain. First, existing studies investigating genotype-phenotype associations have been limited to summed scores of core autism features in smaller sample sizes^19–21^ rather than the underlying latent dimensions. This distinction is important given that autism is phenotypically dissociable^12, 22, 23^ and some associations may emerge only when latent traits are considered. Second, whilst the impact of *de novo* genetic variants on co-occurring developmental disabilities is reasonably well characterised^17, 20, 21^, the impact of common genetic variants is unknown. Third, although sex differences in autism vary by the presence of ID^17, 24, 25^, the sex-differential impact of common genetic variants in autistic individuals with and without ID is unknown. Finally, the impact of latent core autism phenotypes, sex, and *de novo* variants on the common-variant heritability also warrants investigation in large sample sizes.

Here, we address these four questions by combining genetic and phenotypic data from up to 12,893 autistic individuals from four different datasets. We focus on two classes of genetic variants that are robustly associated with autism - *de novo* protein truncating and missense variants in constrained genes (high-impact *de novo* variants)^17, 26^, and polygenic scores, which model the common genetic liability for autism and genetically correlated phenotypes.^16^ Finally, this larger sample size alongside more detailed information on genes underlying severe developmental disorders^27^ also allows us to revisit and provide deeper insights into two additional important issues relevant to heterogeneity in autism: the association of high-impact *de novo* variants with (1) co-occurring developmental disabilities and (2) sex.

## Results

### Identifying latent phenotypes in core autism features

A critical challenge in identifying sources of heterogeneity in autism is understanding the latent structure of core autism phenotypes. Previous studies have conducted genotype-phenotype association studies of core autism features using summed scores of several autistic trait measures and their subscales in relatively modest sample sizes.^19–21^ However, these summed scores may capture multiple aggregated latent traits, introducing statistical noise and limiting interpretability of the results. To this end, we combined two widely used parent-report measures of autistic traits - Repetitive Behaviour Scale - Revised (RBS)^28^ and Social Communication Questionnaire - Lifetime version (SCQ)^29^ - in 24,420 autistic individuals from the Simons Simplex Collection (SSC)^30^ and the SPARK^31^ cohorts. These are the only measures of core autism phenotypes in SPARK, and together capture several social and non-social aspects of the core autism diagnostic domains. We conducted exploratory factor analysis in approximately half of the autistic individuals from SSC (N = 901), randomly selected, followed by confirmatory factor analyses in the remaining half (N = 902) of the SSC and individuals from SPARK (N = 22,617, **Methods**).

We tested 42 different factor models, including bifactor models (**Supplementary Table 1, Supplementary Figure 1**). After multiple iterations, exploratory factor analyses (**Supplementary Figure 2**) identified a correlated six factor model with good theoretical interpretation and confirmatory factor analyses identified fair fit indices (Confirmatory Fit Indices: 0.92 to 0.94; Tucker Lewis Indices: 0.92 to 0.94; Root Mean Square Errors: 0.056 – 0.060). Fit indices increased modestly when including orthogonal method factors in the model (**Supplementary Table 1**). The explained common variances and hierarchical omegas for the bifactor models were low ( < 0.7), suggesting that general factors may not explain the data well (**Supplementary Table 2**). The six identified factors are: 1. Insistence on sameness (F1); 2. Social interaction at age five (F2); 3. Sensory-motor behaviour (F3); 4. Self-injurious behaviour (F4); 5. Idiosyncratic repetitive speech and behaviour (F5); 6. Communication skills (F6) (**Supplementary Table 3**). These broadly correspond to four restricted, repetitive and sensory behaviour factors i.e. non-social factors (Insistence of sameness, Sensory-motor behaviour, Self-injurious behaviour, and Idiosyncratic repetitive speech and behaviour) and two social factors (Social interaction and Communication skills).

All inter-factor correlations were significant and moderate to high in magnitude, with higher correlation among non-social and social factors than between social and non-social factors (**Figure 1A**). Autistic males scored higher (i.e., greater difficulties) than autistic females on all factors except ‘Self-injurious behaviour’ and ‘Insistence on sameness’ (**Figure 1B, Supplementary Table 4a**). All six factors were negatively correlated with full-scale IQ (**Figure 1C**), and this was observed in both males and females separately (**Supplementary Figure 3, Supplementary Table 4b**). In this cross-sectional data, older participants had lower factor scores (i.e. fewer difficulties), with the exception of ‘Social Interaction’ (**Figure 1D**). However, of the 21 items in the ‘Social interaction’ factor, 19 specifically ask about behaviour between ages 4 and 5 (or in the past 12 months for younger children, **Methods**), and this trajectory likely reflects recall bias, as caregivers are likely to report more severe behaviours retrospectively^32^. Similar trends were observed in both males and females (**Supplementary Figure 4**). Some factor scores had modestly higher SNP heritability compared to RBS and SCQ (**Supplementary Table 5**), and there were moderate to high bivariate genetic correlations among the six factors (**Supplementary Table 6**).

**Figure 1:**
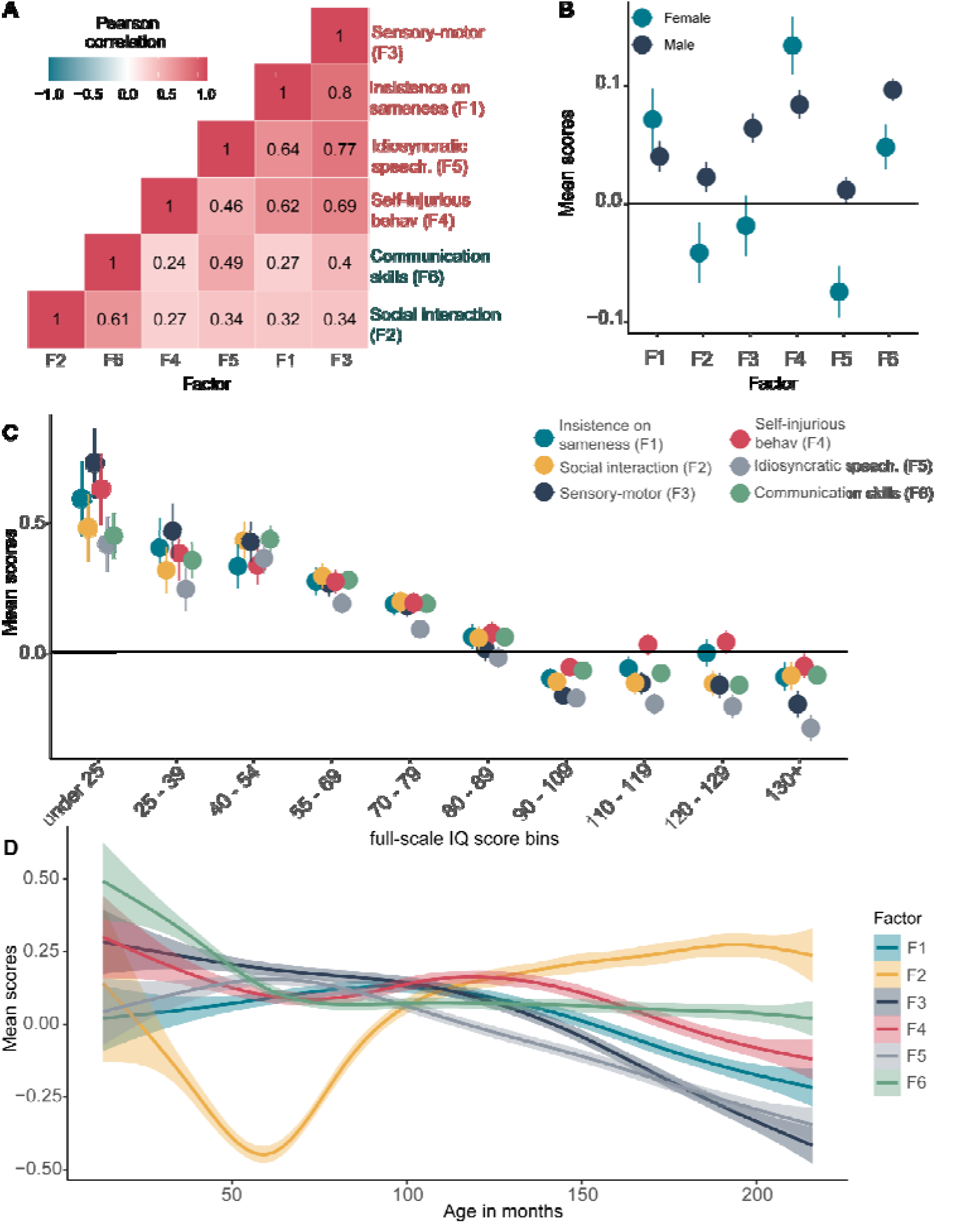
Factor analyses of the core autism features. 1A. Pearson’s Correlation coefficient between the six factors. Factors have been ordered based on hierarchical clustering, demonstrating higher correlations among social (teal labels) and non-social (red labels) factors than between them. 1B. Mean scores and 95% confidence intervals for the six factor scores in males and females. 1C. Mean scores and 95% confidence intervals for the six factor scores in 10 full-scale IQ bins. 1D. Mean factor scores for the six factors across age. The six factors are: 1. Insistence of sameness (F1); 2. Social interaction (F2); 3. Sensory-motor behaviour (F3); 4. Self-injurious behaviour (F4); 5. Idiosyncratic repetitive speech and behaviour (F5); 6. Communication skills (F6). F2 primarily consists of items related to Social interaction at ages 4 – 5 (past 12 months if younger than 4), hence the trajectory likely reflects recall bias in participants.

### Common genetic variants are robustly associated with core autism features but high-impact *de novo* variants are not

Previous smaller scale studies have identified associations with some autism features and both common and *de novo* variants.^19–21^ Here, we expanded both the sample size by combining genetic data from autistic individuals in four cohorts (N_max_ = 12,893), and the number of phenotypes investigated (19 different core and associated features) and conducted genetic association analyses to understand the impact of different classes of genetic variants on these phenotypic features. These 19 features include measures and subscales of measures commonly included for a research diagnosis of autism, parent reports of autistic features, and measures of IQ, adaptive behaviour, and motor coordination (**Methods**).

We first investigated the association between the 19 features and PGS^2#^ for autism, intelligence, educational attainment, and schizophrenia, and, as a negative control, hair colour (N = 2,421 to 12,893, **Supplementary Table 7)**. In multiple regression analyses, autism PGS were associated with increased core autism features (total scores on the RBS and SCQ, and self-injurious factor scores) (**Figure 2A, Supplementary Table 8**), and increased non-verbal IQ. Intelligence PGS were associated with increased full-scale and non-verbal IQ. Educational attainment PGS were associated with increased full-scale and verbal IQ and reduced scores on core autism features. Finally, schizophrenia PGS were associated with reduced adaptive behaviour, measured using the composite score of the Vineland Adaptive Behaviour Scales. The majority of the significant associations (13 out of 16) had concordant effect directions in all cohorts (**Supplementary Figure 5**). We did not identify any significant genotype-phenotype association using hair colour (blonde vs other) as a negative control (**Supplementary Table 8)**.

**Figure 2:**
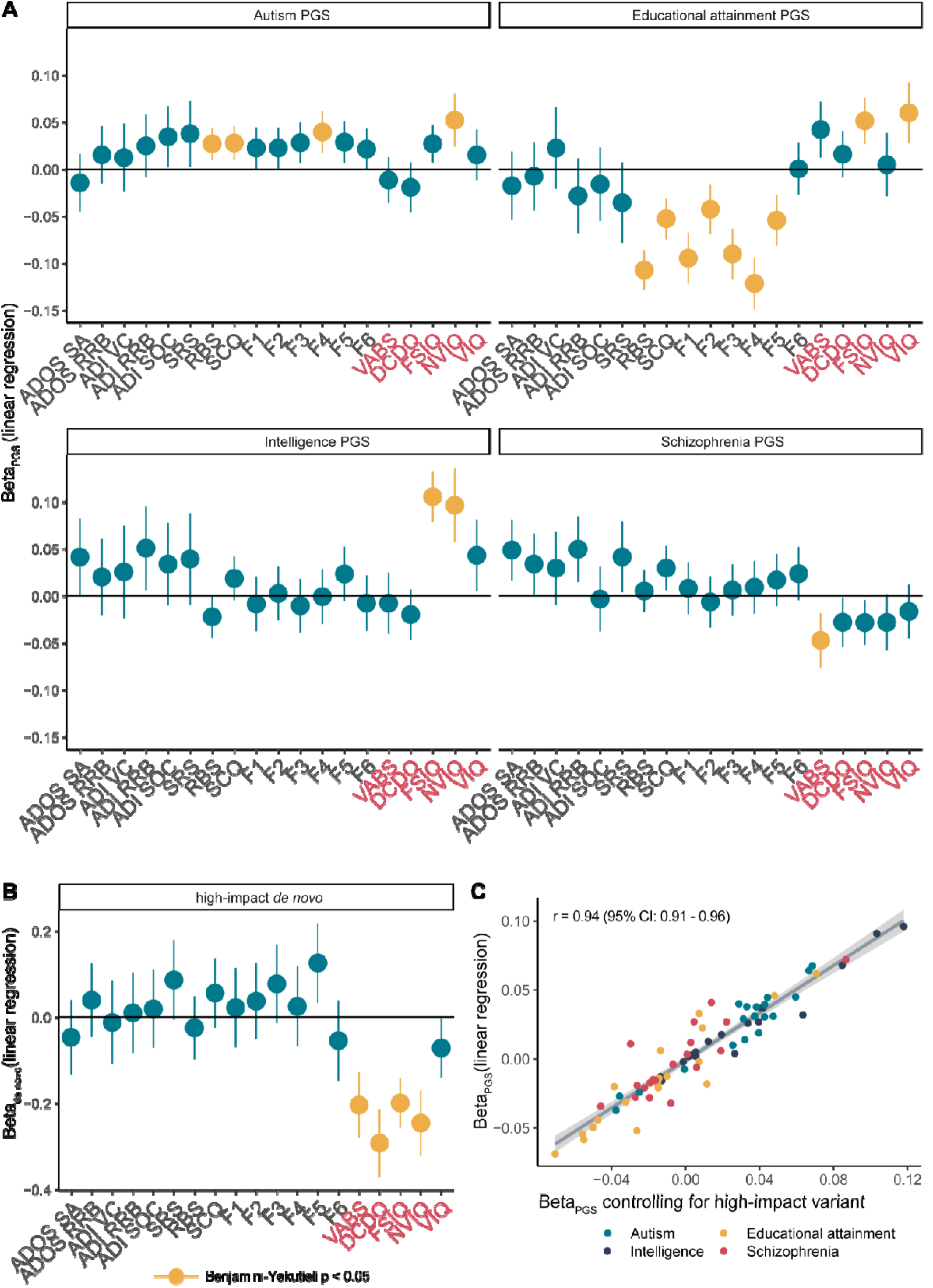
Association of PGS and high-impact *de novo* variants with core and associate autism features. 2A: Associations between the core and associated autism features and PGS for autism educational attainment, intelligence, and schizophrenia. 2B. Associations between high-impact de novo variants and autism features. For all association plots, standardized regression coefficients from linear regressions and 95% confidence intervals are provided. Yellow indicates significant association after Benjamini-Yekutieli correction (corrected p-value < 0.05). Red text indicates associated features, where higher values correspond to greater ability. 2C Correlation between beta coefficients (linear regression) for the four sets of PGS without (y axis) and after (x axis) controlling for the presence of high-impact de novo variants. Phenotypes are: ADOS Social affect (ADOS SA) and restricted and repetitive behaviour (ADOS RRB); ADI verbal communication (ADI VC), social interaction (ADI SOC), and restricted and repetitive behaviour (ADI RRB); Repetitive Behavior Scale-Revised (RBS); Social Communication Questionnaire (SCQ); Insistence of sameness factor (F1); Social interaction factor (F2); Sensory-motor behaviour factor (F3); Self-injurious behaviour factor (F4); idiosyncratic repetitive speech and behaviour (F5); Communication skills factor (F6); Vinelands Adaptive Behaviour Scales (VABS); Development Coordination Disorders Questionnaire (DCDQ); full-Scale IQ (FSIQ); non-verbal IQ (NVIQ); and Verbal IQ (VIQ).

In line with previous results,^17, 20, 21^ the number of high-impact *de novo* variants (protein truncating single nucleotide variants and structural variants, and missense variants with MPC score > 2, N = 2,863 to 4,442) was associated with reduced measures of IQ, adaptive behaviour, and motor coordination. Despite the expanded sample size^17, 20, 21^ and more fine-grained phenotypes^19^ investigated compared to previous analyses, these variants were not robustly associated with any of the core autism features (**Figure 2B, Supplementary Table 9**). The effect sizes of the PGS did not attenuate after controlling for the presence of high impact *de novo* variants (**Figure 2C, Supplementary Table 9,** and **Supplementary Figure 6**), suggesting largely independent effects between common and rare variants.

Measures of IQ are the primary features impacted by both classes of genetic variants, which may reflect both underlying biology and/or our ability to measure IQ with greater reliability than autistic traits. In autistic individuals, full-scale IQ reduced with increasing number of high-impact *de novo* variants but increased with increasing PGS for intelligence (**Figure 3A**). No strong evidence of interaction between PGS for intelligence and high-impact *de novo* variants was observed, suggesting their additive effects on full-scale IQ. Among the significant genotype-phenotype associations, accounting for full-scale IQ did not attenuate the effects of PGS on core autism features (**Figure 3B, Supplementary Table 10**), which was supported by minimal and statistically non-significant genetic correlations between full-scale IQ and the core autism features (**Supplementary Table 6**). This suggests that full-scale IQ does not mediate the association between common genetic variants and core autism features. In contrast, associations between high-impact de novo variants and associated autism features attenuated, partly because of the moderate phenotypic correlations between these features and full-scale IQ (**Figure 4C**).

**Figure 3:**
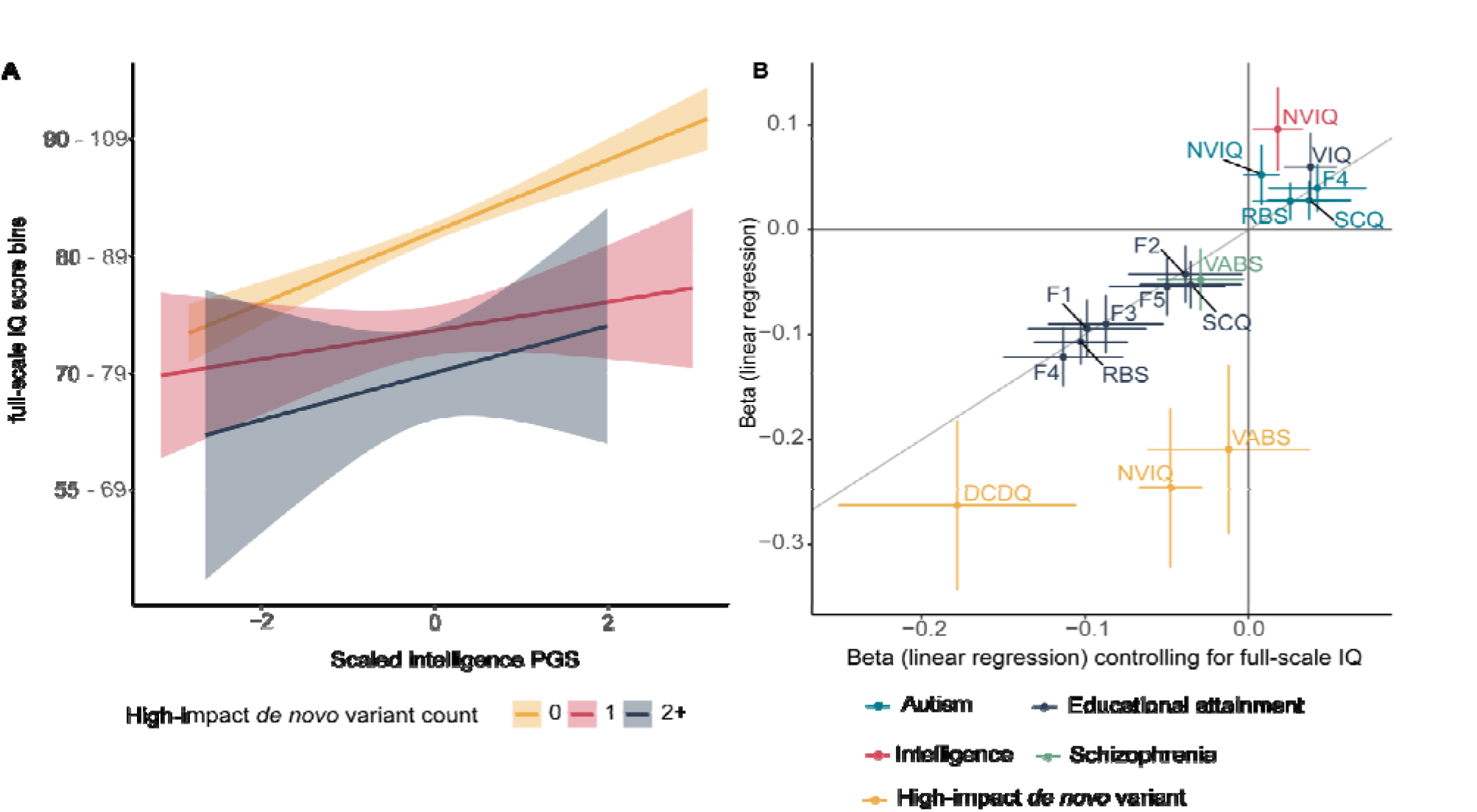
Association between genotype and full-scale IQ, and impact of full-scale IQ on genotype-phenotype associations. 3A: Line plots for full-scale IQ scores as a function of intelligence PGS and counts of high-impact de novo variants in SPARK and SSC (N = 3,197). Only binned full-scale IQ scores were available in SPARK and subsequently, full-scale IQ was binned in SSC and treated as a continuous variable (Methods). 3B: Point estimates of linear regression coefficients for the association between PGS and high-impact de novo variants and core and associated autism features without (y axis) and after (x axis) accounting for full-scale IQ scores. 95% confidence intervals for both regression provided. Only significant genotype-phenotype estimates are plotted. Point estimates closer to the diagonal line indicate no change in beta coefficient (linear regression) after controlling for full-scale IQ. NVIQ = non-verbal IQ, VIQ = Verbal IQ, DCDQ = motor coordination assessed by the Developmental Coordination Disorders Checklist, VABS = adaptive behaviour assessed by the Vineland Adaptive Behaviour Scales, F1 = Insistence on sameness, F2 = Social interaction F3 = Sensory-motor behaviour, F4 = Self-injurious behaviour, F5 = Idiosyncratic repetitive speech and behaviour, F6 = Communication skills, RBS = Repetitive Behaviour Scale, SCQ = Social Communication Questionnaire.

### Mechanisms contributing to core autism phenotypes in autistic individuals with high-impact *de novo* variants

Whilst high-impact variants in some autism-associated genes lead to core autistic features, notably in animal models (e.g.^33, 34^), as a group they were not robustly associated with core autism features (**Figure 2B**). It is unclear if the latent structure of core phenotypes differ in autistic individuals with high-impact *de novo* variants (henceforth, carriers) compared to autistic individuals without any known high-impact de novo variant (henceforth, non-carriers). We thus investigated differences in the latent structure of core autism phenotypes between carriers (N = 325) and non-carriers (N = 2,727). Although likelihood ratio tests identified significant configural invariance violation (i.e., the factor structure dissimilar across groups, p < 2x10^-16^), this was due to the relatively large sample size: the fit indices and visual inspections of the latent structure suggested that the differences were minimal (**Supplementary Table 11**).

Given this, we first investigated whether autistic carriers had higher PGS for autism compared to non-carriers, which may account for core autism features in carriers (additivity). As demonstrated previously, but with a different set of polygenic scores,^19^ autistic carriers had lower PGS for autism than autistic non-carriers (Beta_PGS_ = -0.16, se = 0.045, p = 3.67x10^-4^, linear regression) (**Figure 4A**). This difference was not observed for PGS for educational attainment, IQ, or schizophrenia (**Supplementary Table 12**). However, whilst autistic non-carriers had higher PGS compared to non-autistic siblings (Beta_PGS_ = 0.19, se = 0.023, p = 2.68x10^-15^, logistic regression), autistic carriers (N = 579) were indistinguishable from non-autistic siblings (N = 3,681) based on autism PGS (Beta_PGS_= 0.028, se = 0.045, p = 0.53, logistic regression, **Supplementary Figure 5**).

The PGS in a trio with an affected child can be summarised as the parental mean PGS (henceforth midparental PGS), and the deviation of the affected child’s PGS from the midparental PGS. As previously reported^14^, in this expanded sample size, we identify a overtransmission of autism PGS to autistic individuals (Mean = 0.17, se= 0.01, N = 6,981, p < 2x10^-16^), and curiously, a modest undertransmission to unaffected siblings (Mean = -0.03, se = 0.02, N = 3,832, p = 0.034) (**Figure 4B, Supplementary Table 13**). This likely reflects both reproductive stoppage^35^ and underdiagnosis of autism in the parental generation but greater awareness and opportunity to receive or exclude a diagnosis in children.^36^ Carriers had a modest overtransmission of autism PGS (Mean = 0.08, se = 0.04, N = 579, p = 0.02) whilst this was substantially higher in non-carriers (Mean = 0.18, se = 0.01, N = 4,997, p < 2x10^-16^). Notably, whilst carriers had significantly lower overtransmission compared to non-carriers (p = 0.02), they had a significantly higher overtransmission compared to siblings PGS (p = 9.1x10^-3^), providing additional support for additivity of common and rare genetic variants.

A second hypothesis is that the effect of high-impact *de novo* variants on core autism features is partly mediated by associated autism features. Core and associated autism features are modestly negatively correlated with each other (**Figure 4C**), and given that high-impact *de novo* variants are associated with a relatively sizeable reduction in both full-scale IQ and motor coordination, we reasoned that there will be a knock-on effect on core autism features. The fact that we do not observe a significant association between high-impact *de novo* variants and core autism features (**Figure 2B**) may be due to attenuated correlations between core and associated features in carriers compared to non-carriers^21^. However, tests of matrix correlation equivalence suggested no differences in the phenotypic correlation structures of carriers and non-carriers (p = 9.25x10^-4^, Jennrich test for matrix equivalency). This was supported by the finding of no differences in pairwise Pearson’s correlation coefficients between each of the three associated features and the six factors, SCQ, and RBS between carriers and non-carriers (Fisher’s Z test, all p > 0.05).

An alternate explanation is that we are underpowered to observe this effect. We used simulations to investigate whether we had sufficient statistical power to identify associations between high-impact *de novo* variants and core autism features. Assuming that all effects are completely mediated by only one of the three associated features (full-scale IQ, adaptive behaviour, or motor coordination), power calculations indicate that we have less than 80% power for all core autism features tested (**Figure 4D**). Larger samples may identify significant effects between high-impact *de novo* variants and core autism features, but it will be important to investigate whether the associations are mediated by associated autism features.

**Figure 4:**
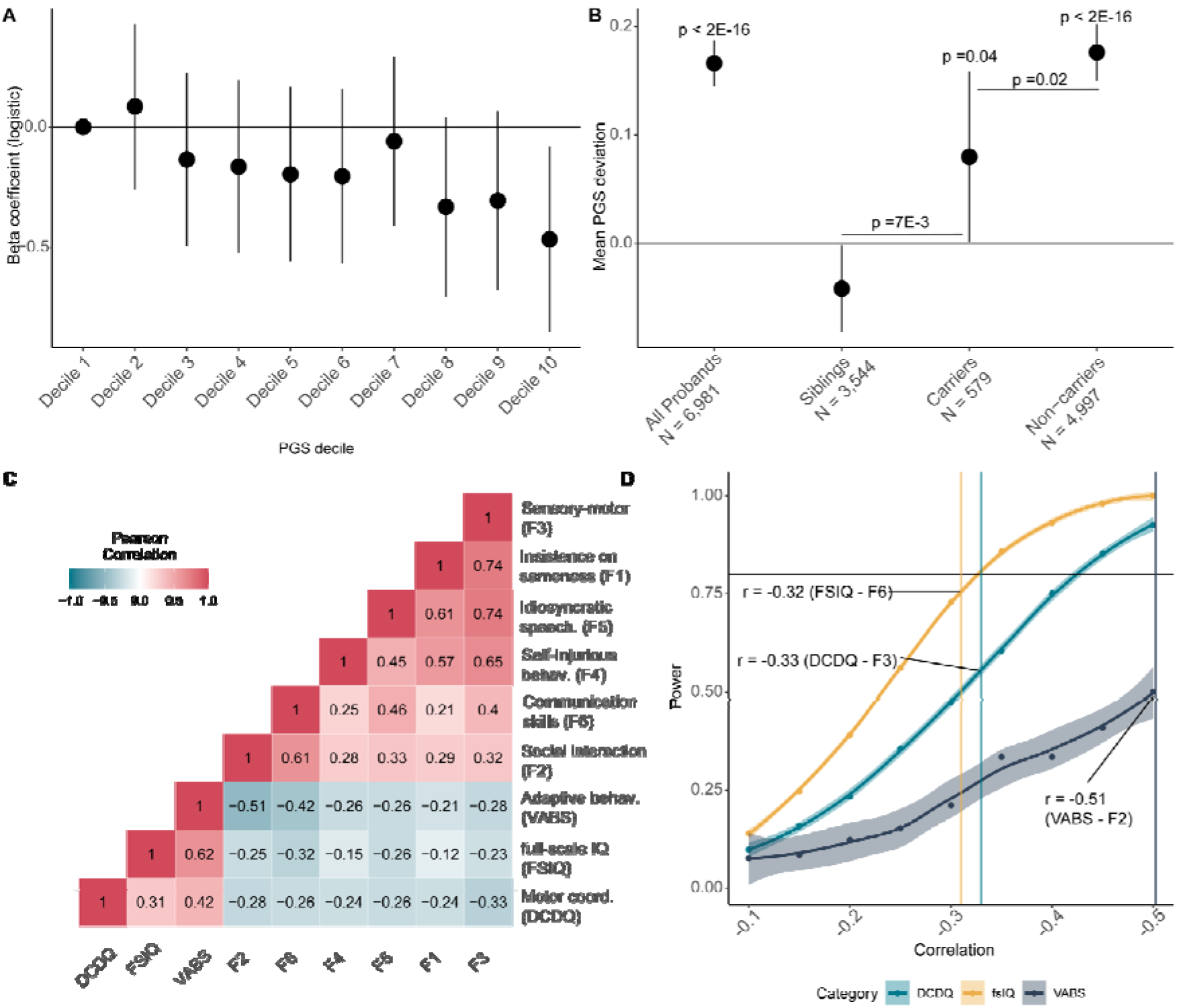
Additivity and impact of high-impact *de novo* variants on core autism features. A. Beta-coefficients (Beta_denovo_) for carrying a high-impact de novo variant per decile of autism PGS in autistic individuals, after accounting for sex, age, 10 genetic principal components, and PGS for educational attainment, intelligence, and schizophrenia, calculated using logistic regression. B. Overtransmission and 95% confidence errors of PGS for autism in all probands, siblings, carriers of high-impact de novo variants, and non-carriers. p-values on top provided for the overtransmission. We also compare differences in overtransmission between carriers and non-carriers and carriers and siblings, and provide the p-values for this from two-tailed Z tests. C. Phenotypic correlation between the core features and associated autism features. D. Statistical power for identifying a significant association between the number of high-impact de novo variants and core features based on the correlation with the three associated features, which is provided in 4C. The highest correlation between a core feature and an associated feature have been indicated on the power graph. DCDQ = motor coordination as measured by the Developmental Coordination Disorders Checklist; fsIQ = full-scale IQ; VABS = adaptive behaviour as measured by the Vineland Adaptive Behaviour Scales.

### Relationship between autism PGS and co-occurring developmental disabilities in autistic individuals

Multiple co-occurring developmental disabilities are commonly observed in autistic individuals, including language, motor, and intellectual developmental disabilities and delays. These represent another source of heterogeneity among autistic individuals. Whilst co-occurring developmental disabilities are associated with high-impact *de novo* variants,^15, 17, 20^ it is unclear whether they are impacted by polygenic scores for autism. We addressed this in SPARK (**Methods**). In line with previous research,^15, 17, 20^ carriers of high-impact *de novo* variants had an increased count of co-occurring developmental disabilities (Beta_denovo_ = 0.31, se = 0.05, p = 1.55x10^-8^, N = 3,089; quasi-poisson regression). In contrast, higher PGS for autism was associated with reduced count of co-occurring developmental disabilities (Beta_PGS_ = -0.037, se = 0.009, p = 3.91x10^-5^, N = 13,435, quasi-poisson regression) even after accounting for the other three PGS (**Figure 5A, Supplementary Table 14a**). Leave-one-out analyses indicated that the results were not driven by any one developmental disability (**Supplementary Figure 8**). Notably, autistic individuals with 5+ co-occurring developmental disabilities did not have statistically higher autism PGS compared to non-autistic siblings (**Figure 5A, Supplementary Table 14b**). In contrast, even when restricting to autistic individuals with no co-occurring developmental disabilities, individuals with a high-impact *de novo* variant were more likely to be autistic compared to non-autistic siblings (**Figure 5A, Supplementary Table 14b**).

The apparent protective effect of autism PGS on co-occurring developmental disabilities has not, to our knowledge, been reported earlier. This can reflect both true protective effects (e.g., PGS for autism increase IQ in both the general population^16, 37^ and in autistic individuals as seen in **Figure 2A**) and the negative correlation between high-impact *de novo* variants and autism PGS. To better delineate this, we investigated the association between the two classes of genetic variants and two well-characterised developmental phenotypes: age of walking independently and age of first words. Both phenotypes were present in autistic individuals from SPARK and SSC, and in siblings from SPARK, allowing us to detect relatively modest effects and draw comparisons with non-autistic siblings. In autistic individuals, autism PGS were associated with earlier age of walking (Beta_PGS_ = - 0.012, se = 0.003, p = 3.2x10^-5^, negative binomial regression) and earlier age of first words (Beta_PGS_ = -0.0125 se = 0.005, p = 0.01, negative binomial regression) whilst high-impact *de novo* variants increased the age for both phenotypes **(Figure 5B, Supplementary Table 15b**). The association between autism PGS and age of walking but not age of first words remained statistically significant after accounting for high-impact *de novo* variants and full-scale IQ (**Supplementary Table 15a**). Similarly, the association between high-impact *de novo* variants and age of walking but not age of first words remained significant after accounting for full-scale IQ (**Supplementary Table 15a**). However, autism PGS were not significantly associated with either age of walking or age of first words in siblings (**Supplementary Table 15a**). Despite the negative association between autism PGS and the two phenotypes, even autistic individuals in the highest decile of autism PGS had higher mean age of walking and age of first words compared to siblings, as did autistic non-carriers (**Figure 5B**) and autistic individuals with no co-occurring developmental disability, suggesting other sources of variation in these phenotypes (**Supplementary Table 15b**).

The above results demonstrate an enrichment of high-impact *de novo* variants in autistic individuals even in the absence of a known co-occurring developmental disability (**Figure 5A**). Yet, there is likely heterogeneity even within the broad class of constrained genes, with differential impact on autism vis-à-vis co-occurring developmental disabilities. Previous research has attempted to disentangle this heterogeneity by comparing counts of disrupting *de novo* variants in autism vs. severe developmental disorders (genetically undiagnosed developmental disorders with accompanying ID and/or developmental delays).^17^ The lack of detailed phenotypic information in the cohorts assessed renders the previous research hard to interpret.^38^ Here we take a different approach to revisit this question. Using the more detailed data on co-occurring developmental disabilities in SPARK, we investigate if constrained genes robustly associated with severe developmental disorders (DD genes)^27^ have differential effects on co-occurring developmental disabilities in autistic individuals compared to other constrained genes (non DD genes). We use the term ‘non DD genes’ for convenience as this list is also likely to contain genes associated with severe developmental disorders which may be discoverable at larger sample sizes but are likely less penetrant (i.e., lower effect size) or lead to increased pre- or perinatal death (i.e., rarer) compared to variants in the DD genes.^27^

In the SPARK cohort, 35.6% of the carriers had high-impact *de novo* variants in DD genes. Autistic individuals were more likely to be carriers of either set of genes compared to non-autistic siblings, which was observed even when restricting to autistic individuals without any known co-occurring developmental disability (**Supplementary Table 14c and d, Figure 5C**). However, whilst the risk for the count of co-occurring developmental disabilities was elevated in carriers of DD genes (Beta_denovo_ = 0.54, se = 0.08, p = 6.48x10^-12^; quasi-poisson regression), this was much more modest for carriers of nonDD genes (Beta_denovo_ = 0.15, se = 0.07, p = 0.035; quasi-poisson regression). Supporting this, autistic carriers of high-impact *de novo* variants in DD genes started walking independently and using words ∼ 3 months later compared to autistic carriers of high-impact *de novo* variants in non-DD genes (p < 0.05 in both, **Figure 5B and Supplementary Table 15b**). These results support a broad phenotypic distinction between the two sets of genes. We ran sensitivity analyses using a larger but overlapping list of genes identified from a highly curated database Developmental Disorder Gene-to-Phenotype (DD2GP)^39^, and identified consistent results (**Supplementary Tables 14 and 15**).

**Figure 5:**
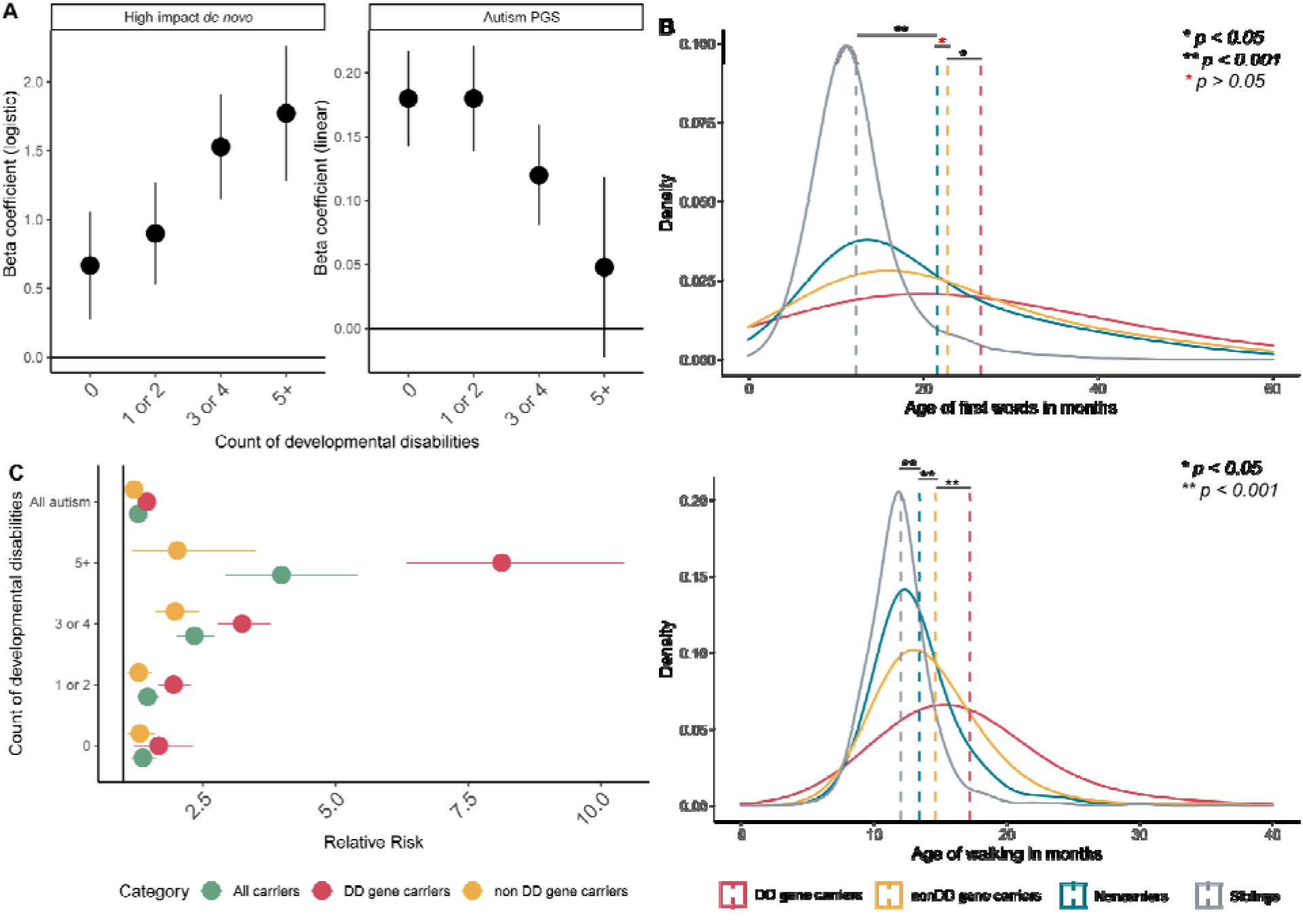
Associations between high-impact *de novo* variants and autism PGS and co-occurring developmental disabilities and delays. A. Beta coefficients for the association of high-impact de novo variants (logistic regression) and autism PGS (linear regression) with case-control status (using sibling controls) by counts of co-occurring developmental disabilities. B. Distribution of and mean age of first words (top) and age of walking (bottom) in siblings, non-carriers, and carriers of high-impact variants in either DD or non-DD genes. p-values were calculated using Wilcoxon Rank Sum Tests. Sample sizes for Figure C are provided in Supplementary Table 14B. C. Relative risk of autism and (any number of) developmental disabilities with 95% confidence intervals for different sets of probands with high-impact de novo variants. Sibling controls were used. All relative risks were statistically significant. All data for Figures A and B are from the SPARK cohort, and sample sizes are provided in Supplementary Table 13.

### Sex-differential overtransmission of autism PGS in autistic individuals without ID

We next turned to another potential source of heterogeneity – sex. Numerous previous studies have found that autistic females are more likely to have high-impact *de novo* variants than autistic males^17, 26, 40, 41^. This observation is thought to support the Female Protective Effect in autism, which suggests that females need a greater genetic liability to cross the autism diagnostic threshold.^13, 40^ However, a similar effect is observed in severe developmental disorders more generally, and is entirely explained by a relatively small number of genes significantly associated with severe developmental disorders (i.e. DD genes).^42^ The observed sex differences in high-impact *de novo* variants in autism may be explained entirely or partly by DD genes, which would suggest a biological mechanism shared by both autism and severe developmental disorders. We thus re-visited sex differences in high-impact *de novo* variants using data from SPARK and SSC (**Supplementary Table 16**). Across all high-impact *de novo variants*, autistic females were more likely to be carriers compared to males (RR = 1.48, 95%CI: 1.27 – 1.71). However, this was explained entirely by high-impact *de novo* variants in DD genes (DD genes: RR = 2.09, 95% CI: 1.73 – 2.54; non-DD genes: RR = 1.14, 95%CI: 0.93 - 1.42) (**Figure 6A**). This sex difference in DD genes remained and did not attenuate after accounting for the total number of co-occurring developmental disabilities in SPARK (Unconditional estimates:Beta_denovo_ = 0.83, se = 0.21, p = 8.15x10^-5^, Conditional estimates: Beta_denovo_ = 0.82, se = 0.22, p = 3.53x10^-4^, logistic regression) and after accounting for full-scale IQ and motor coordination scores in SSC and SPARK (Unconditional estimates: Beta_denovo_ = 1.10, se = 0.15, p = 3.42x10^-13^; Conditional estimates: Beta_denovo_ = 1.31, se = 0.20, p = 8.19x10^-11^, logistic regression). We did not observe sex differences for either gene set in siblings (p > 0.05). These results suggest that sex differences in high-impact *de novo* variants are driven by a relatively small set of highly constrained genes that also increase the likelihood of co-occurring developmental disabilities in autism.

**Figure 6:**
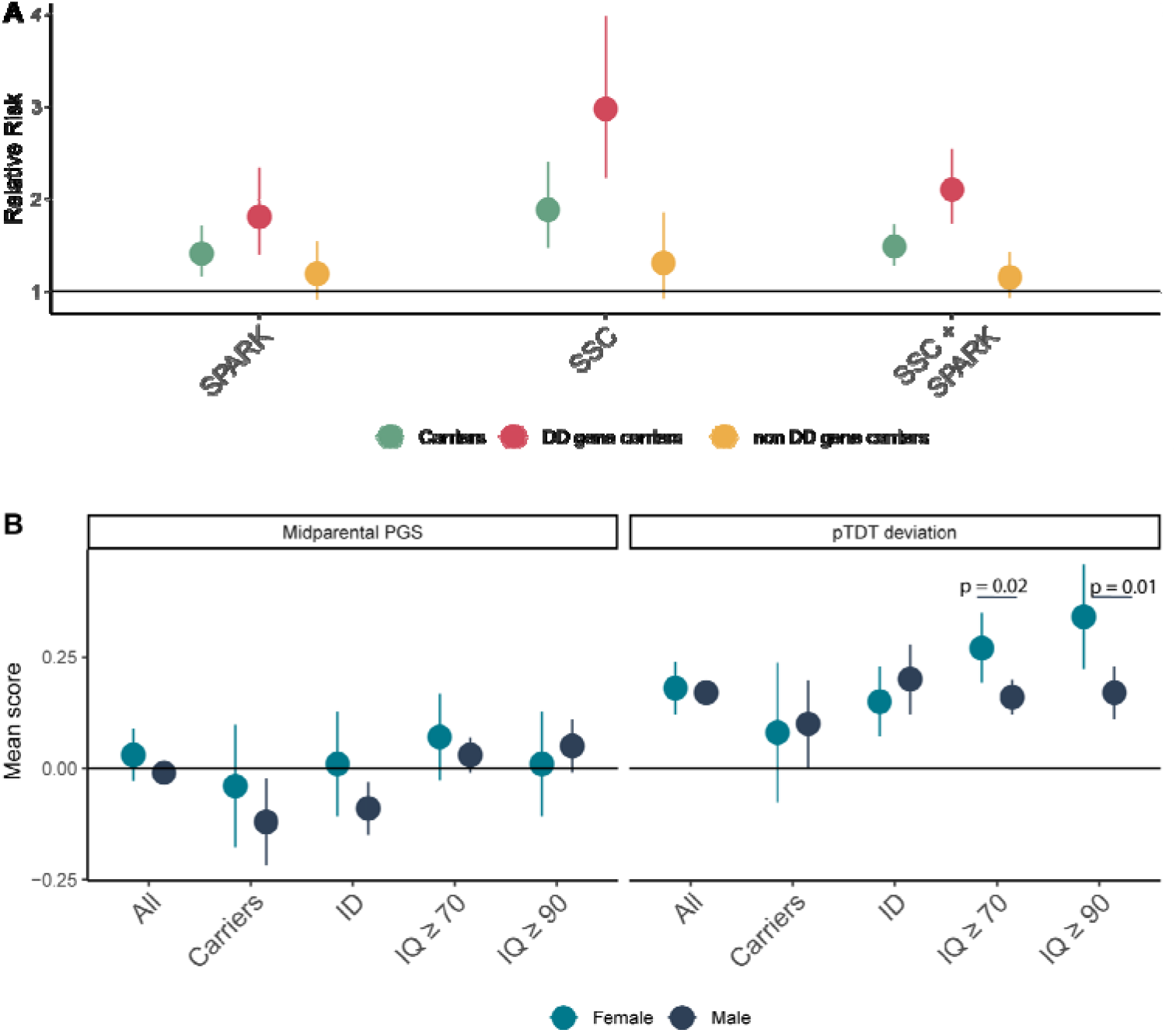
Sex differences in rare and common variants. Figure 6: A: Relative risk and 95% confidence intervals for females compared to males for being a carrier, a DD-gene carrier, and a non DD gene carrier. Sample sizes are provided in Supplementary Table 15. B. Point estimates and 95% confidence intervals showing sex-stratified autism PGS for subgroups of autistic individuals. The first panel provides midparental estimates and the second panel provides the overtransmitted PGS scores. All scores have been standardized to mid-parental means. p-values are provided from two-tailed Z tests. Carriers = carriers of high-impact de novo variants, ID = autistic individuals with co-occurring intellectual disability (fsIQ < 70). Sample sizes are provided in Supplementary Table 17.

Both the contribution of polygenic scores (**Figure 5C)** and the male:female ratio are higher in autistic individuals without ID compared to those with ID, suggesting that polygenic liability for autism may differ between sexes at IQ scores of 70 or above. Recent studies have found higher PGS for autism in females compared to males^19^, and greater overtransmission of PGS for autism in female non-carriers compared to male carriers^43^. Yet, sex differences in polygenic liability have not been investigated while stratifying by the presence of ID. We conducted sex-stratified pTDT to investigate this (N_max_ = 6,981 autistic trios). Whilst PGS for autism were overtransmitted in both male and female probands, this overtransmission did not differ by sex (**Figure 6, Supplementary Table 17**). However, in autistic individuals without ID (IQ>70), females had ∼ 75% higher overtransmission of autism PGS than males (p = 0.02, two-tailed Z test, **Figure 6B**). When using a more conservating IQ threshold of 90 to remove individuals with borderline intellectual functioning, females had double the overtransmission of autism PGS compared to males (Females: Mean = 0.34, se = 0.06, N = 276; Males: Mean = 0.17, se = 0.03, N = 1,328, difference: p = 0.01, two tailed Z-test). We do not find any sex difference in overtransmission for autistic individuals with ID or autistic carriers of a high-impact *de novo* variants. This sex difference in overtransmission was not observed for PGS for educational attainment and intelligence (**Supplementary Table 17**), suggesting that the results are not due to differences in IQ scores between sexes. We also do not find any sex differences in overtransmission of autism PGS in siblings. Furthermore, there was no difference in midparental PGS scores, family income, or parent education by sex or ID (p > 0.05 for all comparisons) - factors correlated with participation in research.^44^ This suggests that these results are unlikely to be explained by sex differences in participation. We cannot, however, distinguish the Female Protective Effect due to common or rare variants from diagnostic bias in the current study.^24, 45^

### Sex and ID but not core autism features impact SNP heritability estimates

Given differences between subgroups of autistic individuals, we next investigated the impact of this heterogeneity on SNP heritability. We used 4,481 unrelated individuals of European ancestries from the ABCD cohort as population controls (**Methods**). Using a rigorous quality control pipeline, we calculated SNP heritability for autism and multiple subgroups (**Figure 7A, Supplementary Table 18**) using two methods - GREML ^46, 47^ and PCGC^48^. All heritability estimates are reported on the liability scale, using sub-group-specific estimates of prevalence (see Methods)

**Figure 7:**
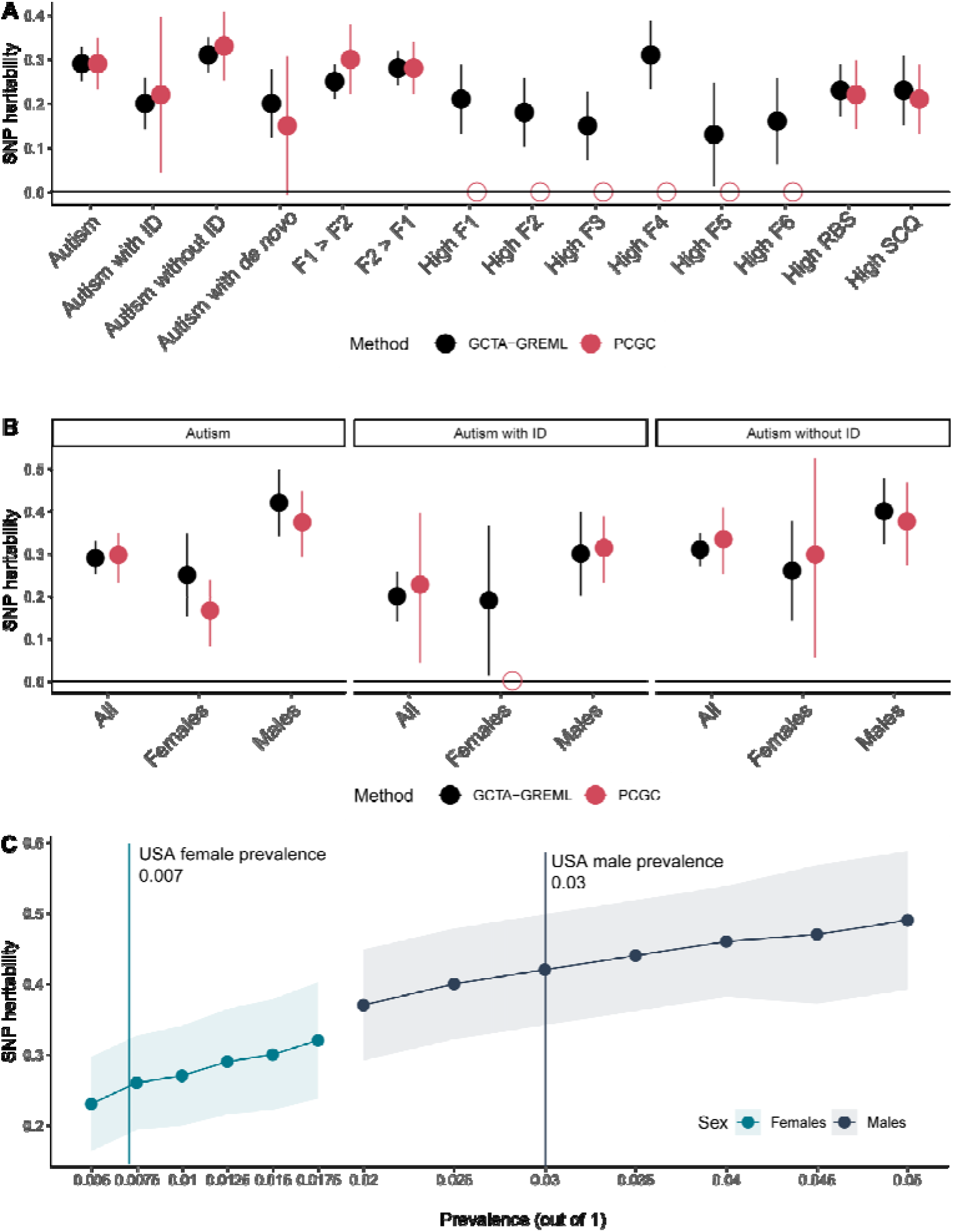
SNP heritability estimates. A: SNP heritability and 95% confidence intervals for various subgroups (males and females combined) of autistic individuals. Estimates from two methods (GCTA-GREML and PCGC) shown. Empty shapes indicate that SNP heritability was not estimated due to low statistical power. B: SNP heritability and 95% confidence intervals for sex- and ID- stratified autism subgroups. Empty shapes indicate that SNP heritability was not estimated due to low statistical power. C: SNP heritability by sex for varying levels of autism prevalence in USA. Shaded regions provide 95% confidence intervals. The six factors are: 1. Insistence of sameness (F1); 2. Social interaction (F2); 3. Sensory-motor behaviour (F3); 4. Self-injurious behaviour (F4); 5. Idiosyncratic repetitive speech and behaviour (F5); 6. Communication skills (F6).

We identified a modest SNP heritability for autism (GCTA: h^2^_SNP_ = 0.29, se = 0.02; PCGC: h^2^_SNP_ = 0.29, se =0.03), which is higher than estimates from iPSYCH^16^ but lower than estimates from the AGRE^49^ and PAGES^50^ cohorts. In line with previous analyses, autistic individuals with ID had lower SNP heritability (GCTA: h^2^_SNP_ = 0.20, se = 0.03; PCGC: h^2^_SNP_ = 0.22, se = 0.09), whilst SNP heritability of autistic individuals without ID was similar to that observed for non-stratified autism (GCTA: h^2^_SNP_ : 0.32, se: 0.02; PCGC: h^2^_SNP_ : 0.33, se: 0.04), but was statistically higher than the SNP heritability of autistic individuals with ID (p = 1.6x10^-3^, two-tailed Z test). SNP heritability for autism in autistic carriers compared to general population controls (agnostic of carrier status) was modest (GCTA: h^2^_SNP_ : 0.20, se: 0.05; PCGC: h^2^ : 0.14, se: 0.08), which is similar to the SNP heritability observed for autistic individuals with ID. However, when comparing autistic high-impact *de novo* carriers with autistic non-carriers, the SNP heritability was not statistically significant (GCTA: h^2^ : 0.14, se: 0.14; PCGC: h^2^ : 0.15, se: 0.19), suggesting that the observed SNP heritability for autistic carriers reflects autism rather than factors associated with the generation of germline mutations.^51, 52^ This result is in line with our pTDT analyses which identify an overtransmission of PGS in carriers, and previous research that has identified a smaller, yet significant heritability for severe developmental disorders.^53^

Stratifying by sex had the largest effect on SNP heritability (**Figure 7B**). Males (GCTA: h^2^_SNP_ : 0.42, se: 0.02; PCGC: h^2^_SNP_ : 0.37, se: 0.04) had approximately 70% higher SNP heritability compared to females (GCTA: h^2^_SNP_: 0.25, se: 0.05, PCGC: h^2^_SNP_: 0.16, se: 0.04; difference: p = 9.3x10^-3^, two-tailed Z test). This difference was observed across a range of prevalence estimates (**Supplementary Table 19, Figure 7C**), after downsampling the number of autistic males to match the number of autistic females (**Supplementary Table 18**), and varying the male-female ratio to 3.3:1 to account for diagnostic bias^45^ (**Supplementary Table 19**). In contrast, stratifying individuals by high scores (1 SD above the mean) on the core autism phenotypes or a combination of two core autism phenotypes modestly reduced or did not alter the SNP heritability for autism (**Supplementary Table 18, Figure 7A)**.

## Discussion

We investigated the genetic correlates of heterogeneity in autism using the largest available genotyped and well-phenotyped autism datasets to date. We make three key observations. First, individual differences among autistic individuals in core and associated features are complex and genetically multifactorial. High-impact *de novo* variants and PGS have differential and often independent effects on these features. This suggests that variants associated with autism as a group can have substantially different effects on presentation. Yet, despite differential effects of PGS on core-autism features, stratifying autism by core autism features has negligible impact on SNP heritability, similar to what has been previously reported at a much smaller scale^18^. In other words, phenotypic homogeneity using only the core autism phenotypes does not increase SNP heritability. This is either because autism is an emergent phenotype that lies at the intersection of multiple phenotypes or because there remains substantial heterogeneity in the underlying cognitive mechanisms that give rise to behavioural differences.

Second, there is additivity between common and high-impact *de novo* variants in autism. These represent the most widely studied class of genetic variants in autism so far, yet emerging evidence suggests a role for other classes (e.g., rare inherited and *de novo* tandem repeats) of genetic variants as well^17, 19, 54, 55^. However, genetic additivity does not imply phenotypic additivity. In other words, the two classes of genetic variants do not have the same effects on either the core or associated autism phenotypes, nor on co-occurring developmental disabilities. Importantly, autism PGS and high-impact *de novo* variants have seemingly opposite effects on co-occurring developmental disabilities. The ‘apparent protective effect’ of autism PGS on co-occurring developmental disabilities reflects both a true protective effect (e.g., for IQ^37^) and the additivity between rare and common variants. Supporting the latter, autism PGS was not significantly associated with earlier age of walking or first words in siblings, though this warrants further investigation in population cohorts. Furthermore, even among high-impact *de novo* variants we identify heterogeneity in effects - genes robustly associated with severe developmental disorders^27^ are the primary drivers of co-occurring developmental disabilities in autism.

Third, we observe sizeable differences in both common and high-impact *de novo* variants based on sex and ID. Females were more likely to have both elevated autism PGS (non-ID only) and have lower SNP heritability, and also more likely to be carriers of high-impact *de novo* variants compared to males, with differential effects based on ID. Whilst this may be interpreted as providing support for the Female Protective Effect^13, 40^, we argue that this is not straightforward. First, the increased burden of high-impact *de novo* variants was observed only with genes associated with severe developmental disorders, not for other constrained genes, despite both sets of genes increasing the liability for autism. This suggests that the Female Protective Effect may be for severe developmental disorders rather than for autism specifically, which warrants further investigation. Second, the higher overtransmission of autism PGS must be interpreted alongside the reduced SNP heritability of autism in females. Assuming high genetic correlation between males and females, reduced SNP heritability in females suggests that higher PGS are required to reach the equivalent levels of genetic liability in males.^56^ Yet, this raises another important question - why do autistic females have a lower SNP heritability compared to autistic males? Does this reflect ascertainment bias in the GWAS cohorts, diagnostic bias, diagnostic overshadowing, camouflaging/masking and/or social stigma?^7, 24, 45^ Several social factors can influence diagnosis in a sex differential manner, and investigating this is paramount to understanding sex differential genetic effects.

Our findings have important implications for using genetics to understand autism. First, genetic investigations without deep phenotyping will certainly advance gene discovery, but will limit both the interpretation and utility of genetics in providing support for autistic individuals who need it. Second, phenotyping needs to be done at scale: effect sizes are modest, and some effects emerge only after considering multiple sources of heterogeneity. This requires concerted efforts from researchers to identify the most relevant phenotypes for investigation, in consultation with the autistic community. Third, both the diagnostic criteria and factors associated with receiving an autism diagnosis are changing^57^, and this will influence our understanding of heterogeneity in autism. Finally, for many autistic individuals, the most disabling aspects are not autism *per se*, but rather co-occurring developmental, physical, and mental health conditions alongside inadequate access to social, educational, financial, and occupational support. Thus, we need to prioritize research that investigates heterogeneity in both autism and co-occurring conditions to develop better targeted support strategies.

## Methods

### Factor analyses

#### Phenotypes and participants

We conducted factor analyses using the Social Communication Questionnaire-Lifetime version (SCQ)^29^ and the Repetitive Behaviour-Revised scale (RBS)^28^. The SCQ, is a widely used caregiver-report of autistic traits capturing primarily social communication difficulties and, to a lesser extent, repetitive and restricted behaviours^29^. There are 40 binary (yes-or-no) questions in total, with the first question focusing on the individual’s ability to use phrases or sentences (total score: 0 to 39). We used the Lifetime version rather than the current version as this was available in both SPARK and SSC. Of note, in the Lifetime version, questions 1 - 19 are about behaviour over the life-time, whilst questions 20 - 40 refer to behaviour between the ages of 4 to 5, or in the last 12 months if the participant is younger. We excluded participants who could not communicate using phrases or sentences (N = 217 in SSC and 17,092 in SPARK) as other questions in the SCQ were not applicable to this group of participants. The RBS is a caregiver-report measure of presence and severity of repetitive behaviours over the last 12 months. It consists of 43 questions assessed on a four-point likert scale (total score: 0 to 129). Higher scores on both measures indicate greater autistic traits.

We restricted our analyses to these two measures as these were the only measures of core autistic traits included in the SSC and SPARK cohorts. Participants had to have completed both measures. We also excluded autistic individuals with incomplete entries in either of the two measures (N = 5,754 only in SPARK). This resulted in 1,803 participants (N = 1,554 males) in SSC, 14,346 (N = 11,440 males) in SPARK version 3 and 8,271 (N = 6,262 males) in extra entries from SPARK version 5 (SSC: Mean age = 108.75, *SD* = 43.29 ; SPARK version 3: Mean age = 112.11 months, SD = 46.43; SPARK version 5: Mean age = months, SD = 48.19). Only the SCQ was available for siblings in SPARK.

#### Exploratory factor analyses

We conducted exploratory factor analysis in a random half of the SSC (N = 901 individuals, of which 782 were males ) using ‘promax’ rotation to identify correlated factors as implemented ‘Psych’^58^ in R. We conducted three sets of exploratory correlated factor analyses: for all items, for social items, for non-social items. Previous studies have provided support for a broad dissociation between social and non-social autism features^12, 23^, and have conducted separate factor analyses of social (e.g.,^59, 60^) and non-social autism features (e.g.^61, 62^). Thus, we reasoned that separating items into social and non-social may aid the identification of covariance structures that may not be apparent when analysing all items together. We divided the data into social (all of SCQ except item 1 and 9 other items and item 28 from RBS) and non-social (9 items from SCQ: Items 8, 11, 12, 14 - 18, and all items from RBS except item 28) items, which was done after discussion between VW and XZ. The ideal number of factors to be extracted was identified from examining the scree plot (**Supplementary Figure 2**), parallel analyses, and theoretical interpretability of the extracted factors. However, we examined all potential models using confirmatory factor analyses as well to obtain fit indices, and the final model was identified using both exploratory and confirmatory factor analyses.

We then applied the model configurations from ‘promax’ rotated exploratory factor analysis for bifactor models, to explore the existence of general factor(s). In addition to a single general factor bifactor model, we divided the data into social and non-social items as mentioned earlier, and conducted bifactor models separately for the social and non-social items. Hierarchical Omega values and explained common variances (ECVs) were then calculated for potential models, as extra indicators of the feasibility of bifactor models, but hierarchical Omega values were not greater than 0.8 for most of the models tested, and ECVs were not greater than 0.7^63–65^ for any of the models tested (**Supplementary Table 2**).

#### Confirmatory factor analyses

Three rounds of confirmatory factor analyses were conducted: first in the second half of the SSC, followed by SPARK participants whose phenotypic data was available in V3 of data release, and then, finally, in SPARK participants whose phenotypic data was available only in V4 or V5 of data release and not in the earlier releases. To evaluate the models, multiple widely adopted fit indices were considered, including the Comparative Fit Index (CFI), Tucker-Lewis Index (TLI), and Root Mean Square Error of Approximation (RMSEA). In CFA, items were assigned only to the factor with the highest loading to attain parsimony. We conducted three broad sets of confirmatory factor analyses: (1) confirmatory factor analyses of all correlated factor models; (2) confirmatory factor analyses of the autism bifactor model; (3) confirmatory factor analyses of social and non-social bifactor models. For each of these confirmatory factor models we limited the number of factors tested based on the slope of the scree plots, and based on the number of items loading onto the factor (five or higher). For the confirmatory factor analyses of social and non-social bifactor models we iteratively combined various numbers of social and non-social group factors. In bifactor models, items without loading onto the general factor in the correspondent EFA were excluded. Items were allocated to different group factors which were identified based on the highest loading (items with loading < 0.3 were excluded). Due to the ordinal nature of the data, all CFAs were conducted using the Diagonally Weighted Least Squares estimator (to account for the ordinal nature of the data) in the R package Lavaan 0.6-5^66^. We identified the model most appropriate for the data at hand with TLI and CFI > 0.9 (TLI and CFI > 0.95 for bifactor models), low RMSEA, and good theoretical interpretability based on discussions between authors VW and XZ. Additionally, as sensitivity analyses, the identified model (correlated six-factor model) was run again with two orthogonal method factors mapping onto SCQ and RBS-R, to investigate if the fit indices remained high after accounting for covariance between items derived from the same measure, as these measures vary subtly on the period of time evaluated. We also re-analysed the identified model and after removing items loading onto multiple factors (> 0.3) to provide clearer theoretical interpretation of the model. For genetic analyses, we used factor scores from the correlated six-factor model without including the orthogonal method factors and without dropping the multi-loaded items.

### Genetic quality control

#### Participants

We conducted analyses using data from four cohorts of autistic individuals: The Simons Simplex Collection (SSC, N = 8,813 samples)^30^, the Autism Genetic Resource Exchange (AGRE, CHOP sample) (N_max_ = 1,200 autistic individuals)^67^, the AIMS-2-TRIALS LEAP sample (N_max_ = 262 autistic individuals)^68^, and SPARK (N = 29,782 samples)^31^. For sibling comparisons, we included siblings from SSC (N = 1,829) and SPARK (N = 12,260). For trio-based analyses, we restricted to complete trios in SSC (N = 2,234) and SPARK (N = 4,747). For all analyses we restricted the sample to autistic individuals who passed genetic quality control and who had phenotypic information.

#### Genetic quality control

Quality control was conducted for each cohort separately, by array. We excluded participants with genotyping rate < 95%, excessive heterozygosity (±3 standard deviations from the mean), non-European ancestry as detailed below, mismatched genetic and reported sex, and, for families, Mendelian errors > 10%. SNPs were excluded with genotyping rate < 10%, or if they deviated from Hardy-Weinberg equilibrium (p < 1x10^-6^). Given the ancestral diversity in the SPARK cohort, Hardy-Weinberg equilibrium and heterozygosity were calculated in each genetically homogeneous population separately. Genetically homogeneous populations (corresponding to five super populations – Africa, East Asian, South Asian, Admixed American, and European) were identified using the 5 genetic principal components (PCs) calculated using SPARK and 1000 genomes Phase 3 populations^69^, and clustered using UMAP^70^. PCs were calculated using LD-pruned SNPs (r^2^ = 0.1, window size = 1000 kb, step size = 500 variants, after removing regions with complex LD patterns) using GENESIS^71^, which accounts for relatedness between individuals, calculated using KING^72^.

Imputation was conducted using Michigan Imputation Server^73^ using the 1000 genomes Phase 3 v5 as the reference panel^49^ (for AGRE and SSC), using HRC r1.1 2016 reference panel^74^ (for AIMS-2-TRIALS), or using TOPMED imputation panel^75^ (for both releases of SPARK). Details of imputation have been previously reported^76^. SNPs were excluded from polygenic risk scores if they had minor allele frequency < 1%, had an imputation r^2^ < 0.4, or were multiallelic.

#### Polygenic scores

We restricted our polygenic score (PGS) associations to four GWAS. First, we included a GWAS of autism from the latest release from the iPSYCH cohort (iPSYCH-2015). This includes 19,870 autistic individuals and 39,078 individuals without an autism diagnosis. GWAS was conducted on individuals of European ancestry, with the first 10 genetic principal components included as covariates using a logistic regression as provided in Plink. Further details are provided elsewhere.^43^ We additionally included GWAS for Educational attainment (N = 766,345, excluding the 23andMe dataset)^77^, 2. Intelligence (N = 269,867)^78^, and 3. Schizophrenia (69369 cases and 236,642 controls)^79^. These GWAS were selected given the relatively large sample size and modest genetic correlation with autism. Additionally, as a negative control, we included PGS generated from a GWAS of hair colour (blonde vs other, N = 43,319 blondes and 342,284 others) from the UK Biobank, which was downloaded from here: <u>Genome wide association study ATLAS (ctglab.nl).</u> This phenotype has comparable SNP heritability to the other GWAS used (0.15, se = 0.014), is unlikely to be genetically or phenotypically correlated with autism and related traits, and has a large enough sample size to be a reasonably well-powered negative control.

PGS were generated for three phenotypes using Polygenic Risk Scoring using continuous shrinkage (PRS-CS)^80^, which is among the best-performing polygenic scoring methods using summary statistics in terms of variance explained^81^. In addition, this method bypasses the step of having to identify a p-value threshold. We set the global shrinkage prior (phi) to 1E-02, as is recommended for highly polygenic traits. Details of the SNPs included are provided in **Supplementary Table 3**.

*de novo* variants were obtained from Antaki et al., 2021^19^. *de novo* variants (structural variants and single nucleotide variants) were called for all of the SSC samples and a subset of the SPARK samples (Phase 1 genotype release, single nucleotide variants only). To identify high-impact *de novo* SNVs, we restricted data to variants with a known effect on protein. These are damaging variants: “transcript_ablation”, “splice_acceptor_variant”, “splice_donor_variant”, “stop_gained”, “frameshift_variant”, “stop_loss”, “start_loss”, or missense variants with MPC^82^ (Missense Badness, PolyPhen-2, and Constraint) scores >2. We further restricted data to variants in constrained genes with a LOEUF score < 0.37^83^, which represents the topmost decile of constrained genes. For SVs, we restricted data to SVs affecting the most constrained genes i.e., LOEUF score < 0.37, representing the first decile of most constrained genes. We did not make a distinction between deletions or duplications. To identify carriers, non-carriers, and parents, we restricted our data to samples in SPARK and SSC who had been exome-sequenced, and families in which both parents and the autistic proband(s) passed the genotyping QC.

For genes associated with severe developmental disorders, we obtained the list of constrained genes that are significant genes associated with severe developmental disorders from Kaplanis et al., 2020^27^. To investigate the association of this set of genes with autism and developmental disorders, we first identified autistic carriers with a high-impact *de novo* variant and then divided this group into carriers who had at least one high-impact *de novo* variant in a DDD gene, and carriers with high-impact *de novo* variants in other constrained genes.

Only individuals with undiagnosed developmental disorders are recruited into the Deciphering Developmental Disorders study, and as such known genes associated with developmental disorders that are easy for clinicians to recognise and diagnose may be omitted from the genes identified by Kaplanis et al., 2020.^27^ To account for this bias, we ran sensitivity analyses using a larger but overlapping list of genes identified from the highly Developmental Disorder Gene-to-Phenotype database (DDG2P). From this database, we used constrained genes that are either ‘confirmed’ or ‘probable’ developmental disorder genes and genes where heterozygous variants lead to developmental phenotypes (i.e. monoallelic or X-linked dominant).

### Phenotypes

#### Core and associated autism features

We identified 19 autism core and associated features which: 1. Are widely used in studies related to autism; 2. Are a combination of parent-, self-, other-report and performance-based measures to investigate if reporter status affects the PGS association, 3. Are collected in all three cohorts; and 4. Cover a range of core and associated features in autism. The core features are:

1. ADOS^84^: Social Affect (ADOS SA)
2. ADOS^84^: Restricted and Repetitive Behaviour domain total score (ADOS RRB)
3. ADI^85^: Communication (verbal) domain total score (ADI VC)
4. ADI^85^: Restricted and repetitive behaviour domain total score (ADI RRB)
5. ADI^85^: Social domain total score (ADI SOC)
6. Repetitive Behaviour Scale – Revised^28^ (RBS)
7. Parent-reported Social Responsiveness Scale - 2^86^: Total raw scores (SRS)
8. Social Communication Questionnaire (SCQ)^29^
9. Insistence of sameness factor (F1)
10. Social interaction factor (F2)
11. Sensory-motor behaviour factor(F3)
12. Self-injurious behaviour factor (F4)
13. Idiosyncratic repetitive speech and behaviour (F5)
14. Communication skills factor (F6)

The associated features are :

1. Vineland Adaptive Behaviour Scales^87^: Composite standard scores (VABS)
2. Full-scale IQ (fsIQ)
3. Verbal IQ (vIQ)
4. Non-verbal IQ (nvIQ)
5. Developmental Coordination Disorders Questionnaire^88^ (DCDQ)

Measures of IQ were quantified using multiple methods across the range of IQ scores in AGRE, SSC, and LEAP. In SPARK, IQ scores were available based on parent reports on ten IQ score bins (see: **Figure 1C**). We used these as full-scale scores. For analyses involving SPARK and SSC, we converted full-scale scores from the SSC into IQ bins to match what is available in SPARK, and treated it as a continuous variable based on examination of the frequency histogram (**Supplementary Figure 9**). For the six factors, we excluded individuals who were minimally verbal (see **Methods** section on Factor analyses), but these individuals were not excluded for analyses with other autism features.

#### Developmental phenotypes

We identified seven questions relating to developmental delay in the SPARK medical screening questionnaire. These are all binary questions (yes/no). Summed scores ranged from 0 to 7. The developmental phenotypes include presence of:

1. Intellectual disability, cognitive impairment, global developmental delay, or borderline intellectual functioning
2. Language delay or language disorder
3. Learning disability (LD, learning disorder, including reading, written expression, math, or NVLD (Nonverbal learning disability))
4. Motor delay (e.g., delay in walking) or developmental coordination disorder
5. Mutism
6. Social (Pragmatic) Communication Disorder (as included in DSM IV TR and earlier)
7. Speech articulation problems

We included age of first words and age of walking independently for further analyses. This was recorded using parent-report questionnaires in SPARK and in ADI-R^85^ in SSC. Whilst other developmental phenotypes are available, we focussed on these two as these represent major milestones in motor and language development and are relatively well-characterized.

### Statistical analyses

#### Genetic association analyses

For each cohort, PGS and high-impact *de novo* variants were regressed against the autism features with sex and the first 10 genetic principal components as covariates in all analyses, with all continuous independent variables standardised. In addition, array was included as a covariate in SSC and AGRE datasets. This was using linear regression for standardised quantitative phenotypes, logistic regression for binary phenotypes (e.g association between PGS and presence of a high-impact *de novo* variant), poisson regression for count data (number of developmental disorders/delays, not standardised), and negative-binomial regression for age of walking independently/age of first words, not standardised (MASS^89^ package in R).

For the association between genetic variables and core and associated autism phenotypes, we first conducted linear regression analyses for the four PGS first using multivariate regression analyses using data from SPARK (waves 1 and 2), SSC, AGRE, and AIMS-2-TRIALS LEAP. This is of the form:

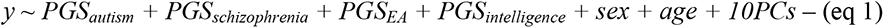

For negative control, we added the negative control as an additional independent variable in equation 1

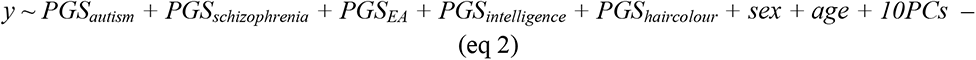

In AGRE and SPARK we ran equivalent mixed-effects models with family ID modelled as random intercepts to account for relatedness between individuals. This was done using the lme4^90^ package in R.

For high-impact *de novo* variants, we included the count of high-impact *de novo* variants as an additional independent variable in equation 1 and ran regression analyses in SPARK (Wave 1 only) and SSC. To ensure interpretability across analyses, we retained only individuals who passed the genotypic QC, which included only individuals of European ancestries. Family ID was included as a random intercept:

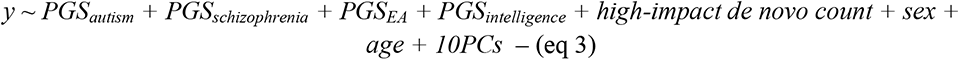

Effect sizes were meta-analysed across the three cohorts using inverse variance weighted meta-analyses with the following formula:

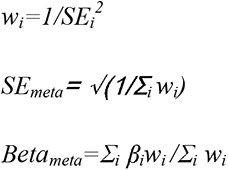

Where *β_i_* is the standardized regression coefficient of the PGS, *SE_i_* is the associated standard error, and *w_i_* is the weight. p-values were calculated from Z scores. Given the high correlation between the autism features and phenotypes, we used Benjamini-Yekutieli False Discovery Rates to correct for multiple testing (corrected p value < 0.05).

In SPARK and SSC, we investigated the association between PGS (equation 1) and being a carrier of a high-impact *de novo* variant (equation 3) and the age of first walking and first words using negative binomial regression, and conducted inverse variance meta-analyses (equation 4). We ran the same analyses in SPARK to investigate the association between PGS (equation 1) and high-impact *de novo* variants (equation 3) and counts of co-occurring developmental disabilities (quasi-poisson regression). Leave one out analyses were conducted by systematically excluding one of seven co-occurring developmental disabilities and reconducting the analyses.

To investigate additivity between common and high-impact *de novo* variants, we conducted logistic regression with carrier status as a dependent binary variable and all PGS included as independent variables and genetic PCs, sex, and age included as covariates. This was done separately in SPARK (Wave 1) and SSC and meta-analysed as outlined earlier.

#### Phenotypic analyses

Statistical significance of differences in factor scores between sexes were computed using t-tests. Associations with age and IQ bins were conducted using linear regressions after including sex as a covariate.

Matrix equivalency tests were conducted using the Jennrich Test in the Psych^58^ package in R. Power calculations were conducted using simulations. Statistical differences between pairwise correlation coefficients (carriers vs non-carriers) in core and associated features were tested using the package cocor^91^ in R. Using scaled existing data on full-scale IQ, adaptive behaviour, and motor coordination, we generated correlated simulated variables at a range of correlation coefficients to reflect the correlation between the six core factors and the three associated features. We then ran regression analyses using the simulated variable and high-impact de novo variants as provided in equation 3. We repeated this a thousand times and counted the fraction of outcomes where the association between high-impact *de novo* variant count and the simulated variable had p < 0.05 to obtain statistical power. Differences in age of walking and age of first words between groups of autistic individuals and siblings were calculated using Wilcoxon Rank-Sum tests.

#### Sex differences: Polygenic transmission disequilibrium tests

Polygenic transmission deviation was conducted using polygenic Transmission Disequilibrium Tests^14^. To allow comparisons with midparental scores, residuals of the autism PGS were obtained after regressing out the first 10 genetic principal components.

These residuals were standardized by using the parental mean and standard deviations. We obtained similar results using PGS that have not been residualized for the first 10 genetic principal components. We defined individuals without co-occurring intellectual disability (ID) as individuals whose full-scale IQ is above 70 in SSC and SPARK. Additionally, in SPARK, we excluded any of these participants who had a co-occurring diagnosis of “Intellectual disability, cognitive impairment, global developmental delay, or borderline intellectual functioning”. Analyses were conducted separately in the SSC and SPARK cohorts and meta-analysed using inverse-variance weighted meta-analyses. We additionally conducted pTDT analyses in non-autistic siblings to investigate differences between males and females.

#### Sex differences: high-impact de novo variants

For sex differences in high-impact de novo variants, we calculated relative risk in autistic females vs males based on: 1. All carriers; 2. Carriers of DD genes; and 3. Carriers of non-DD genes (SPARK wave 1 and SSC). For sensitivity analyses, we conducted logistic regression with sex as the depedent variable and carrier status for DD genes, and either full-scale IQ and motor coordination scores (in SPARK wave 1 and SSC) or number of developmental disorders (only in SPARK wave 1) as covariates. For each sensitivity analysis we provide the estimates of the unconditional analysis as well (i.e. without the covariates).

### Heritability analyses

We opted to conduct heritability analyses using unscreened population controls rather than family controls (i.e., pseudocontrols or unaffected family members), as this likely reduces SNP heritability^92^ owing to parents having higher genetic liability for autism compared to unselected population controls^49^ and due to assortative mating^93^. Case-control heritability analyses were conducted using the ABCD cohort as population controls; specifically, the ABCD child cohort in the USA, recruited at the age of 9 or 10. They are reasonably representative of the US population in terms of demographics and ancestry. As such they represent an excellent comparison cohort for the SPARK and SSC cohorts. ABCD was genotyped using the Smokescreen^TM^ genotype array, a bespoke array designed for the study containing over 300,000 SNPs. Genetic quality control was conducted identical to SPARK. Genetically homogeneous groups were identified using the first five genetic principal components followed by UMAP clustering with the 1000 Genomes data. We restricted our analyses to 4,481 individuals of non-Finnish European ancestries in ABCD. Scripts for this are available here: https://github.com/vwarrier/ABCD_geneticQC. Imputation was conducted, similar to SPARK, using the TOPMED imputation panel.

For case-control heritability analyses we combined genotype data from ABCD, and from autistic individuals from SPARK and SSC. We restricted to 6,328,651 well-imputed SNPs (r^2^ > 0.9) with a minor allele frequency > 1% in all datasets. Furthermore, we excluded multi-allelic SNPs, and SNPs with minor allele frequency difference of > 5% between the three datasets, and, in the combined dataset, were not in HWE (p > 1x10^-6^) or had a genotyping rate < 99%. We additionally excluded related individuals as identified using GCTA-GREML and individuals with genotyping rate < 95%. We calculated genetic PCs for the combined dataset using 52,007 SNPs with minimal linkage disequilibrium (r^2^ = 0.1, 1000 kb, step size of 500 variants, removing regions with complex long-range LD). Visual inspection of the PC plots did not identify any outliers (**Supplementary Figure 10**). Whilst our quality control procedure is stringent, we note that there will be unaccounted for effects in the SNP heritability due to fine-scale population stratification, differences in genotyping array, and participation bias in the autism cohorts. However, our focus is on the differences in SNP heritability between subgroups of autistic individuals and unaccounted for case-control differences will not affect this.

We calculated SNP heritability for autism, and additionally in subgroups stratified for the presence of ID, sex, sex and ID together, and presence of high-impact de novo variants. We also conducted SNP heritability in subgroups of autistic individuals with scores > 1SD from the mean for each of the six factors, autistic individuals with F1 scores > F2, and autistic individuals with F2 scores > F1.

We calculated observed scale SNP heritability (baseline and subgroups) using GCTA-GREML^46, 47^ and additionally, using Phenotype Correlation-Genotype Correlation (PCGC)^48^. In all models except for the sex-stratified models, we included sex, age in months, and the first 10 genetic principal components as covariates. In the sex-stratified models we included age in months and the first 10 genetic principal components as covariates. For sex-stratified heritability analyses, both cases and controls were from the same sex. For GCTA-GREML, observed scale SNP heritability was converted into liability scale SNP heritability using equation 23 in Lee et al., 2011^94^. PCGC estimates SNP heritability directly on the liability scale using the prevalence rates from Maenner et al., 2020^95^. For all analyses we ensured that the number of cases does not exceed the number of controls, with a maximum case:control ratio of 1.

We used prevalence rates from Maenner et al., 2020^95^ which provides prevalence of autism among 8 year olds (1.8%). The study also provides prevalence rates by sex and by the presence of intellectual disability. However, there is wide variation in autism prevalence. We thus re-calculated the SNP heritability across a range of state-specific prevalence estimates obtained from Maenner et al., 2020^95^. For estimates of liability-scale heritability for subtypes defined by factor scores > 1 SD from the mean, we estimated a prevalence of 16% of the total prevalence. For F1 > F2, and F2 > F1, prevalence was estimated at 50% of the total autism prevalence. Estimating approximate population prevalence of autistic individuals with high-impact *de novo* variant carriers is difficult due to ascertainment bias in existing autism cohorts. However, a previous study has demonstrated that the mutation rate for rare protein truncating variants is similar between autistic individuals and siblings from the SSC and autistic individuals and population controls from the iPSYCH sample in Denmark, which does not have a participation bias,^96^ implying that the *de novo* mutation rate in autistic individuals from SPARK and SSC may be generalisable. Using sex-specific proportion of *de novo* variant carriers and autism prevalence, we calculated a prevalence of 0.2% for being an autistic carrier of a high-impact *de novo* variant.

For sex-stratified SNP heritability analyses, we additionally calculated SNP heritability for a range of state-specific prevalence estimates to better model state-specific factors that contribute to autism diagnosis. In addition, using a total prevalence of 1.8%, we estimated SNP heritability using a male-female ratio of 3.3:1^45^ to account for diagnostic bias that may inflate the ratio.

We used GCTA-GREML to also estimate SNP heritability for the six factors, full-scale IQ, and the bivariate genetic correlation between them. We used the same set of SNPs used in the case-control analyses. We were unable to conduct bivariate genetic correlation for the case-control datasets due to limitations of sample size.

### Ethics

We received ethical approval to access and analyse de-identified genetic and phenotypic data from the three cohorts from the University of Cambridge Human Biology Research Ethics Committee.

### Data availability

Genetic and phenotypic data for SFARI and SPARK are available upon application and approval from the Simons Foundation (<u>SFARI | Autism Cohorts</u>). Data for AGRE is available upon application and approval from Autism Speaks (<u>AGRE - Autism Genetic Resource Exchange | Autism Speaks</u>). Data for EU-AIMS Leap is available upon application and approval to the EU-AIMS LEAP committee (<u>The LEAP Study (eu-aims.eu)</u>).

### Scripts

All scripts used in this study are available here:

- Genetic QC and imputation in SSC: vwarrier/SSC_liftover_imputation: Basic scripts used for imputing the SSC genotyped datasets (github.com)
- Genetic QC and imputation in SPARK: vwarrier/SPARK_QC_imputation: QC and imputation of the SPARK dataset (github.com)
- Genetic QC and imputation in ABCD: <u>vwarrier/ABCD_geneticQC (github.com)</u>
- Bespoke genetic analyses: vwarrier/autism_heterogeneity: This git has the code for the heterogeneity in autism project (github.com)

We used the following software packages:

- PRScs: getian107/PRScs: Polygenic prediction via continuous shrinkage priors (github.com)
- TOPMED imputation server: TOPMed Imputation Server (nih.gov)
- Plink: PLINK 2.0 (cog-genomics.org)
- GCTA-GREML: PLINK 2.0 (cnsgenomics.com)
- PCGC: PCGC Regression | dougspeed.com

## Data Availability

Data availability
Genetic and phenotypic data for SFARI and SPARK are available upon application and approval from the Simons Foundation (SFARI | Autism Cohorts). Data for AGRE is available upon application and approval from Autism Speaks (AGRE - Autism Genetic Resource Exchange | Autism Speaks). Data for EU-AIMS Leap is available upon application and approval to the EU-AIMS LEAP committee (The LEAP Study (eu-aims.eu)).

## Acknowledgements

V.W. is funded by St. Catharine’s College, Cambridge. This study was funded by grants to SBC from the Medical Research Council, the Wellcome Trust, the Autism Research Trust, the Templeton World Charity Foundation, and to T.B. from the Institut Pasteur, the CNRS, The Bettencourt-Schueller and the Cognacq-Jay Foundations, the APHP and the Université de Paris. SBC was funded by the Autism Research Trust, the Wellcome Trust, the Templeton World Charitable Foundation, and the NIHR Biomedical Research Centre in Cambridge, during the period of this work. The Medical Research Council (MRC) funded the Cambridge Autism Research Database (CARD) that made this study possible. SBC also received funding from the Innovative Medicines Initiative 2 Joint Undertaking (JU) under grant agreement No 777394. The JU receives support from the European Union’s Horizon 2020 research and innovation programme and EFPIA and AUTISM SPEAKS, Autistica, SFARI. His research was also supported by the National Institute of Health Research (NIHR) Applied Research Collaboration East of England (ARC EoE) programme. T.M.M. is supported by U.S. National Institutes of Mental Health (NIMH) grant MH117014. The views expressed are those of the authors, and not necessarily those of the NIHR, NHS or Department of Health and Social Care. We acknowledge with gratitude the generous support of Drs Dennis and Mireille Gillings in strengthening the collaboration between S.B.-C. and T.B., and between Cambridge University and the Institut Pasteur. The views expressed are those of the author(s) and not necessarily those of the NHS, the NIHR or the Department of Health. The AIMS-2-TRIALS LEAP receives support from the European Union’s Horizon 2020 research and innovation programme and EFPIA and AUTISM SPEAKS, Autistica, SFARI. A full list of the authors and affiliations in the AIMS-2-TRIALS LEAP group is provided in the Supplementary Information. The iPSYCH team was supported by grants from the Lundbeck Foundation (R102-A9118, R155-2014-1724 and R248-2017-2003), NIMH (1U01MH109514-01 to ADB) and the universities and university hospitals of Aarhus and Copenhagen. The Danish National Biobank resource was supported by the Novo Nordisk Foundation. High-performance computer capacity for handling and statistical analysis of iPSYCH data on the GenomeDK HPC facility was provided by the Center for Genomics and Personalized Medicine and the Centre for Integrative Sequencing, iSEQ, Aarhus University, Denmark (grant to ADB). We thank Jonathan Sebat for sharing the *de novo* variant calls in the SPARK and SSC datasets.

---

**Supplementary Figure 1:**
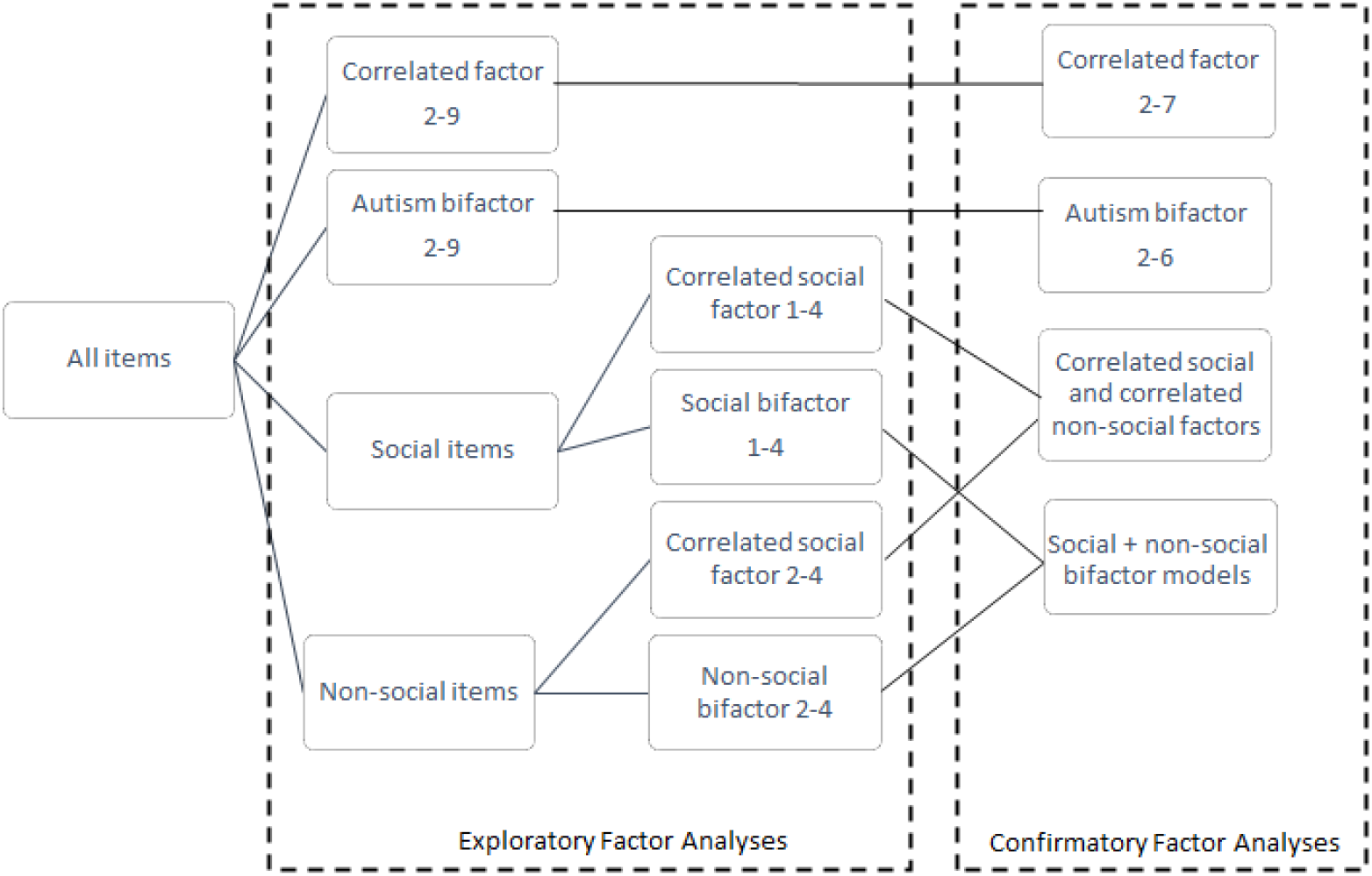
Flowchart of the factor models tested. We tested 42 models in total using a series of exploratory and confirmatory factor analyses. This includes two sets of correlated factor models (correlated factor, and correlated social and correlated non-social factors), and two sets of bifactor models (Autism bifactor and social + non-social bifactor models).

**Supplementary Figure 2:**
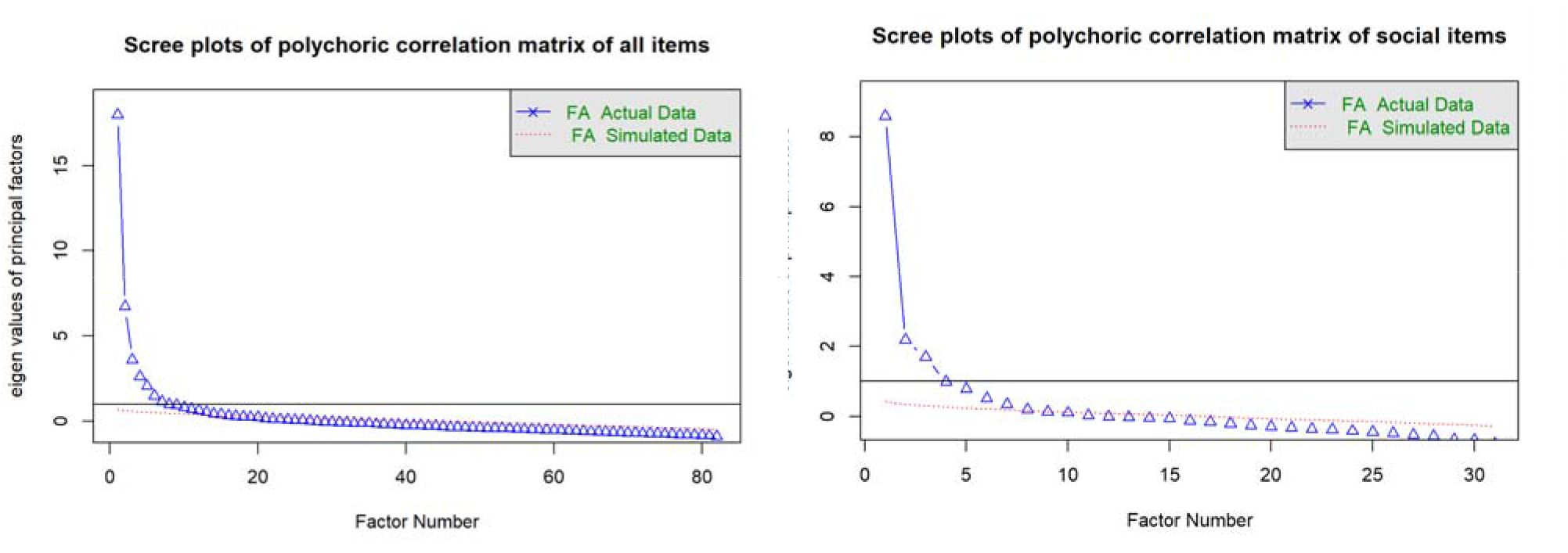
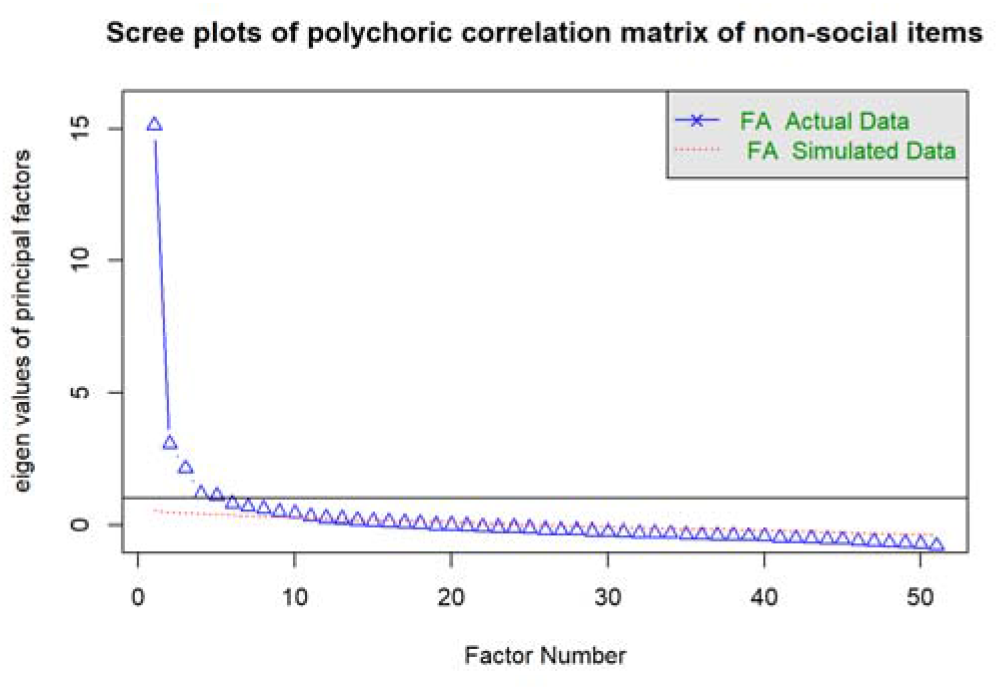
Scree plots for the exploratory factor models. Scree plots for: (1) Correlated factor; (2) Correlated social factor; and (3) Correlated non-social factor models. Examination of the scree plot suggested 6 correlated factors, 2 social factors and 4 non-social factors.

**Supplementary Figure 3:**
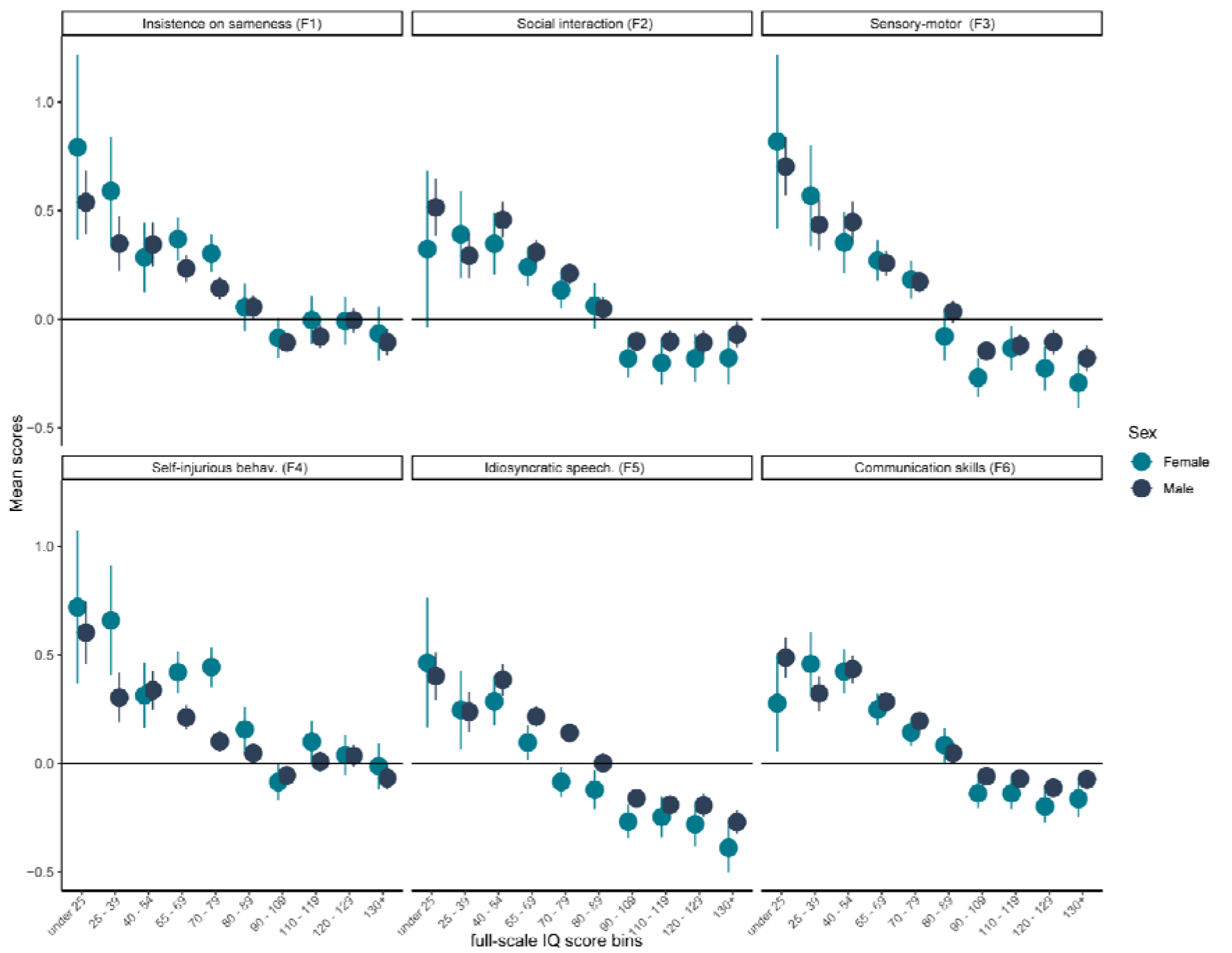
Factor scores by sex and full-scale IQ bins. Mean scores and 95% confidence intervals for the six factor scores in 10 full-scale IQ bins, stratified by sex

**Supplementary Figure 4:**
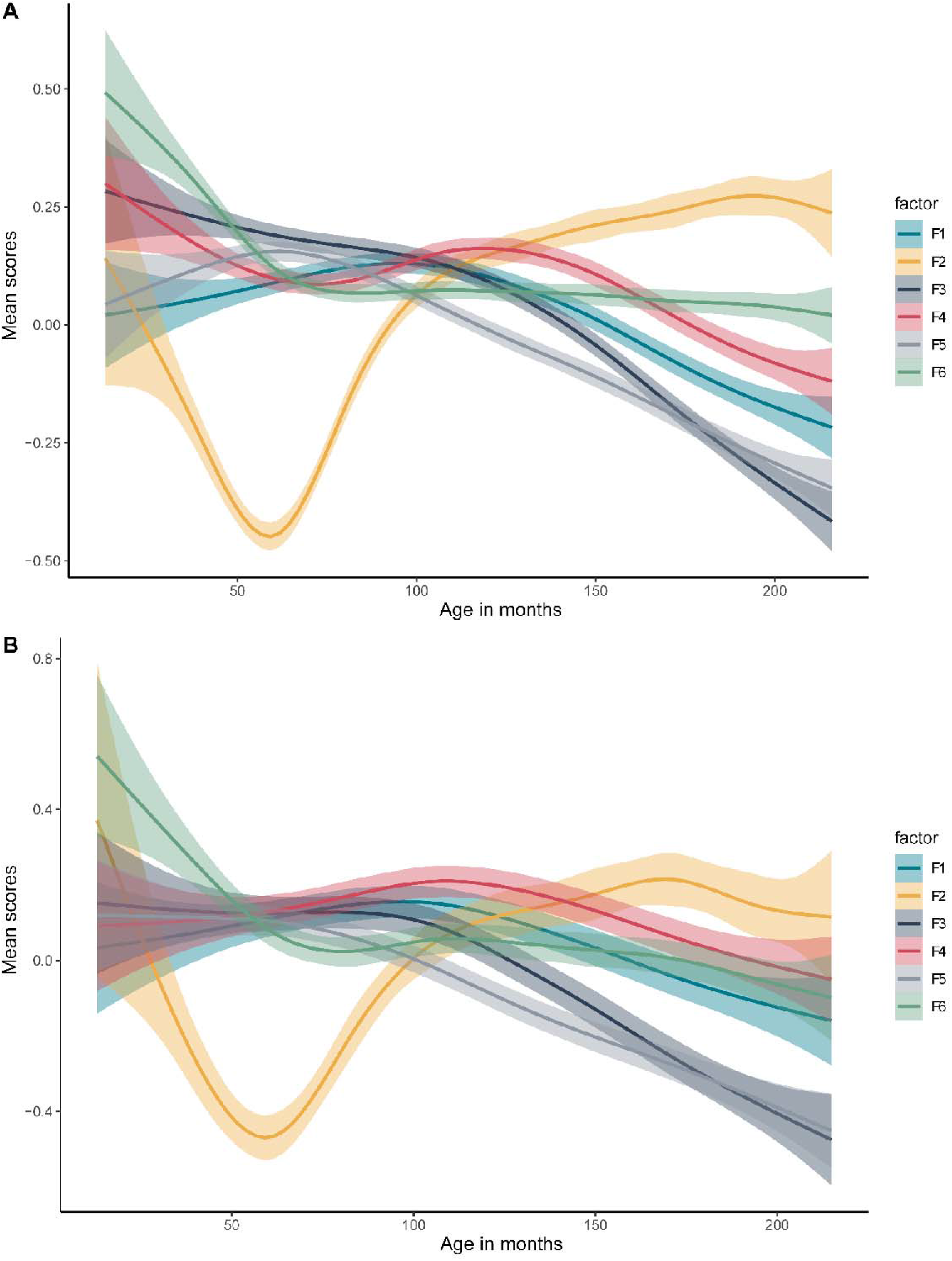
Age related trajectories in factor scores. A. Age related trajectories in males. B Age related trajectories in females. The six factors are: 1. Insistence on sameness (F1); 2. Social interaction (F2); 3. Sensory-motor behaviour (F3); 4. Self-injurious behaviour (F4); 5. Idiosyncratic repetitive speech and behaviour (F5); 6. Communication skills (F6). F2 primarily consists of items related to Social interaction at ages 4 – 5, hence the trajectory likely reflects recall bias in participants.

**Supplementary Figure 5:**
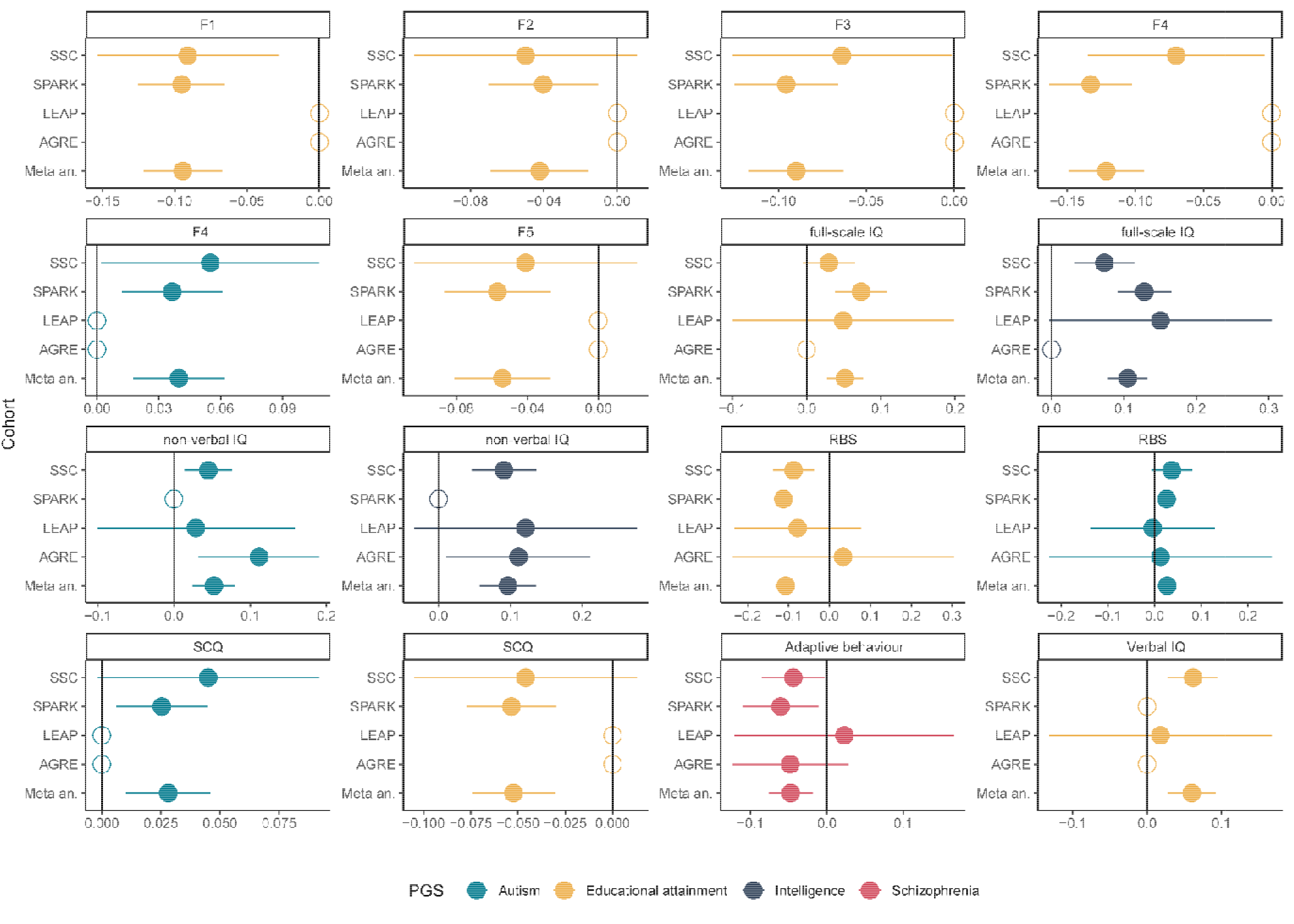
Effect directions for the significant PGS associations. Regression beta and 95% confidence intervals for the significant PGS-feature associations by cohort. Empty circles represent cohorts where the phenotypes were not available (F1, F2, F3, F4, F5, and SCQ were not available in AGRE and LEAP; full-scale IQ and verbal IQ were not available in AGRE, non-verbal IQ and verbal IQ were not available in SPARK). Meta an. Represent the meta-analysed estimates and associated 95% confidence intervals. Adaptive behaviour was measured using the composite scores from the Vinelands Adaptive Behaviour Scales. The five factors are: 1. Insistence on sameness (F1); 2. Social interaction (F2); 3. Sensory-motor behaviour (F3); 4. Self-injurious behaviour (F4); 5. idiosyncratic repetitive speech and behaviour (F5).

**Supplementary Figure 6:**
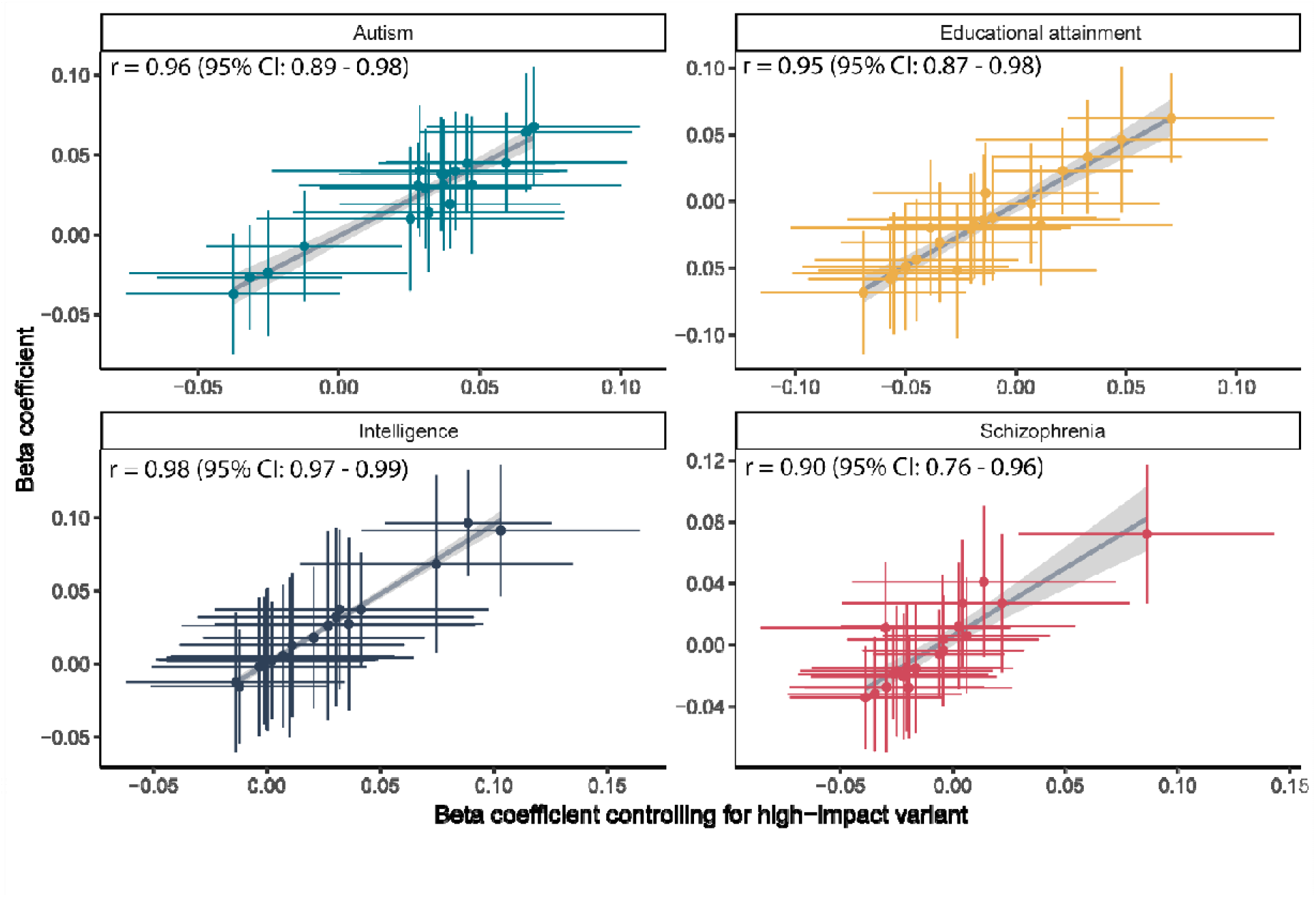
Correlation between beta coefficients for the four sets of PGS without and after controlling for the presence of high-impact *de novo* variants. Correlation coefficients and associated 95% confidence intervals are provided in the inset.

**Supplementary Figure 7:**
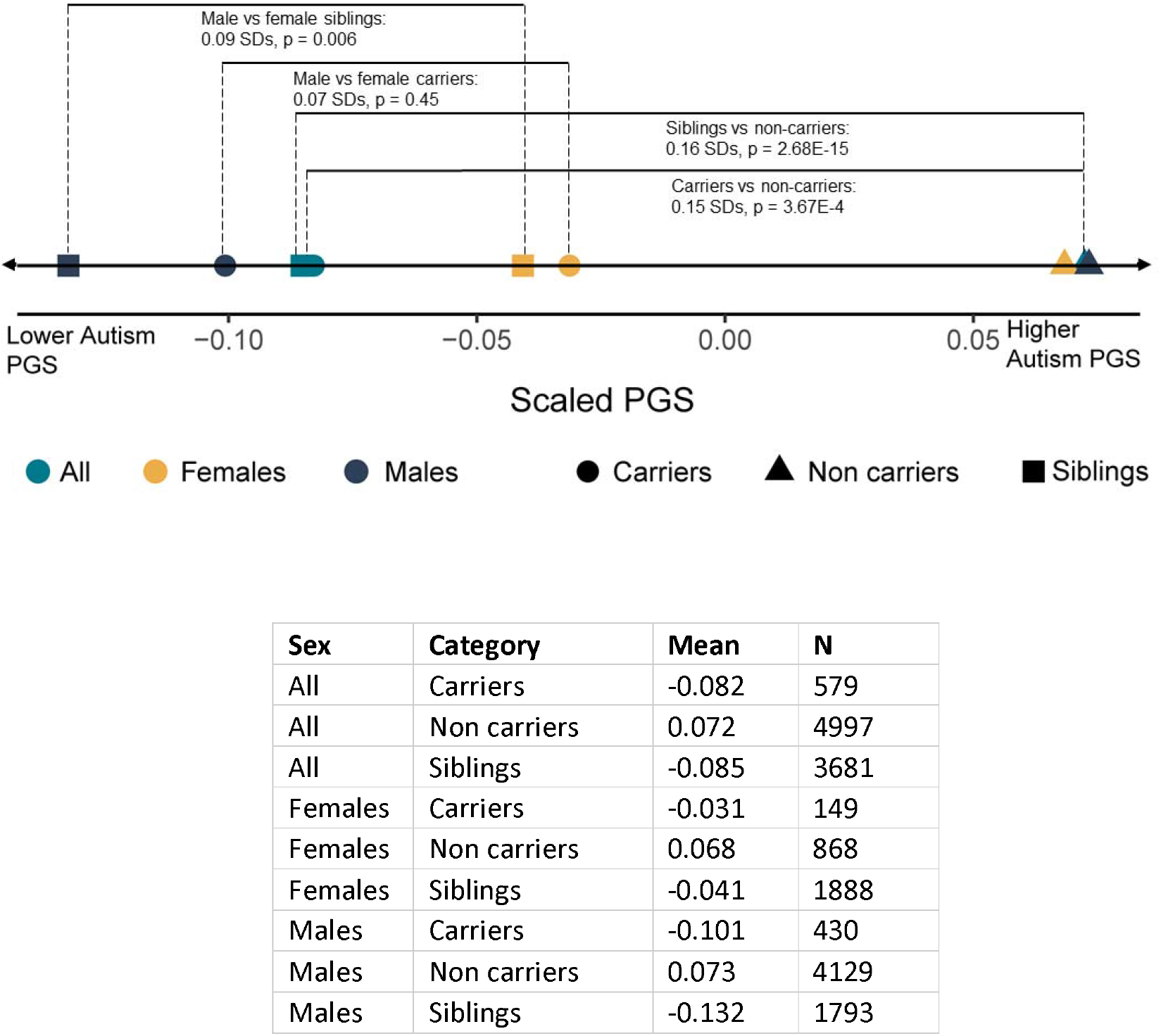
Differences in autism PGS by sex, and diagnostic and carrier status. Differences in standardised autism PGS (mean = 0, standard deviation = 1). Line is drawn to scale. Standard deviations (SDs) and p-values have been provided for select comparisons where visual inspection of the plot identified sizable differences in PGS between groups. p-values have been calculated using linear regression using autism PGS residualised for 10 genetic principal components, and with sex (non-stratified comparisons only) and cohort included as covariates. The table provides the mean PGS and sample size for each group.

**Supplementary Figure 8:**
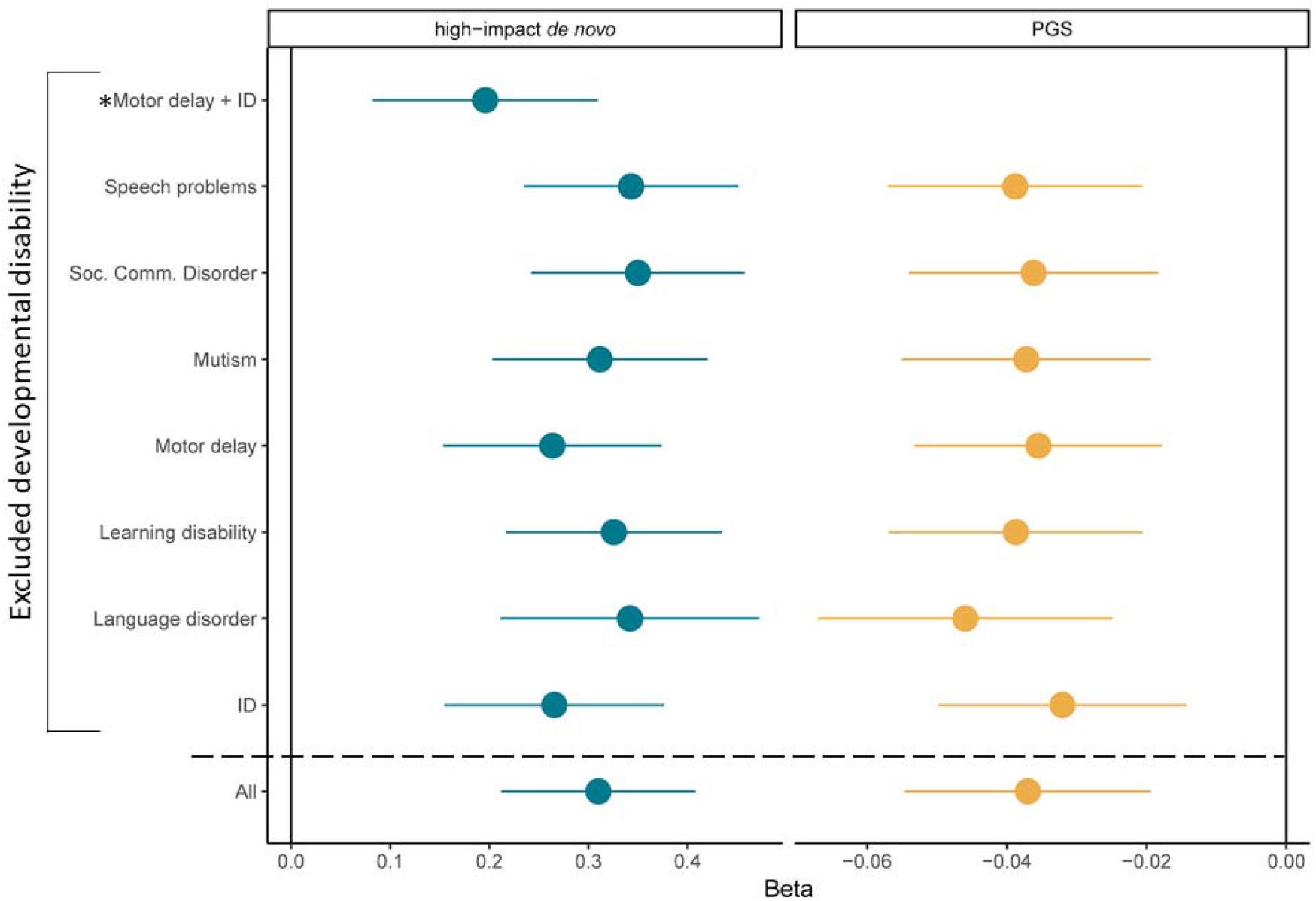
Leave-one-out analyses for genotype-developmental disability associations. Leave-one-out analyses for the associations between autism polygenic scores or high-impact de novo variants and count of developmental disabilities. Regression betas and 95% confidence intervals provided. For high-impact de novo variants, we additionally conducted leave-one-out analyses after excluding both motor delay and ID (indicated using *), given the associations between high-impact variants and both IQ and motor coordination. We also provide the beta and 95% confidence intervals for the original regression for the count of all seven developmental disabilities (‘All’) for comparison.

**Supplementary Figure 9:**
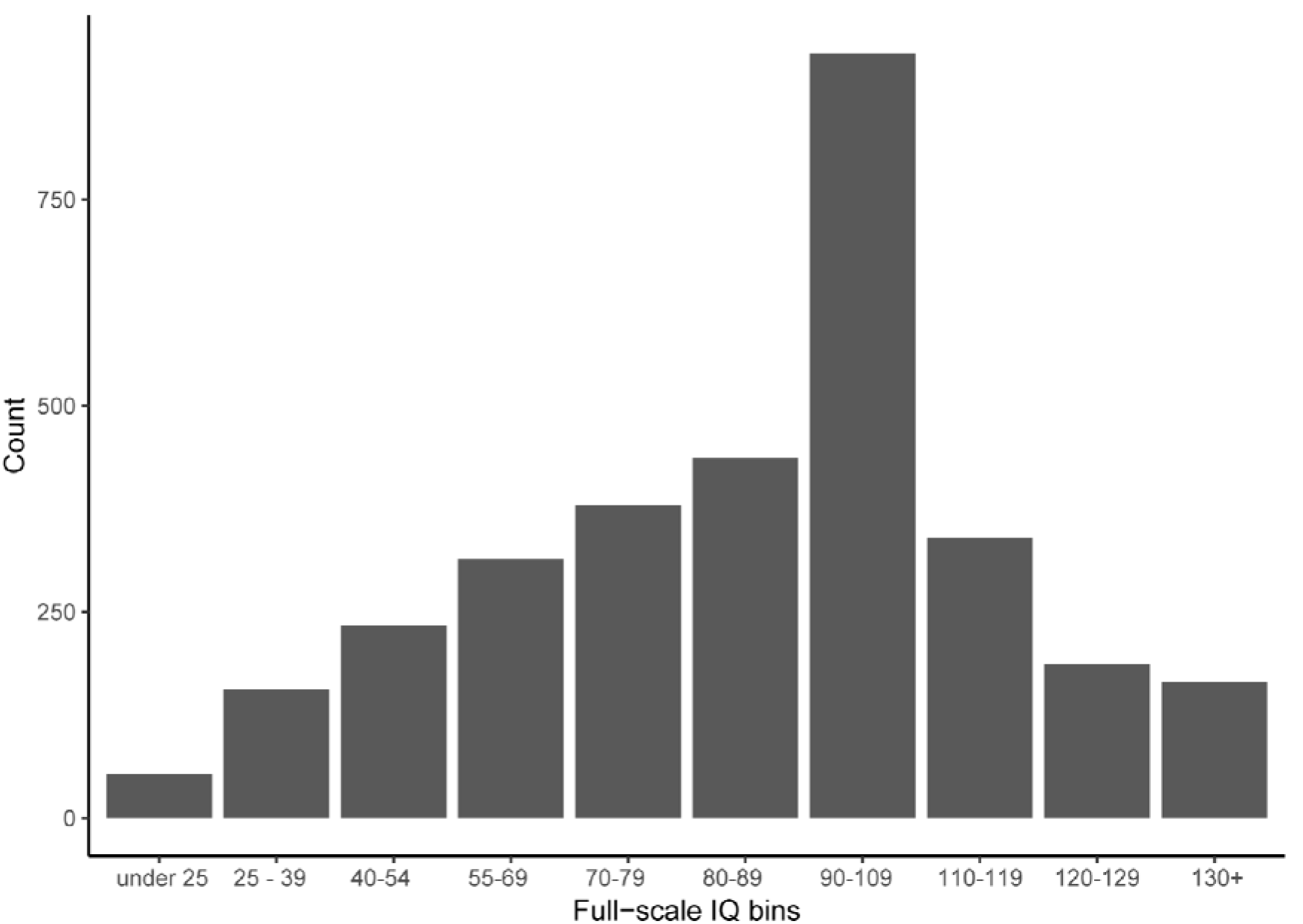
Distribution of full-scale IQ bins in SPARK and SSC combined. Frequency histogram of binned full-scale IQ scores from the SPARK and SSC cohorts.

**Supplementary Figure 10:**
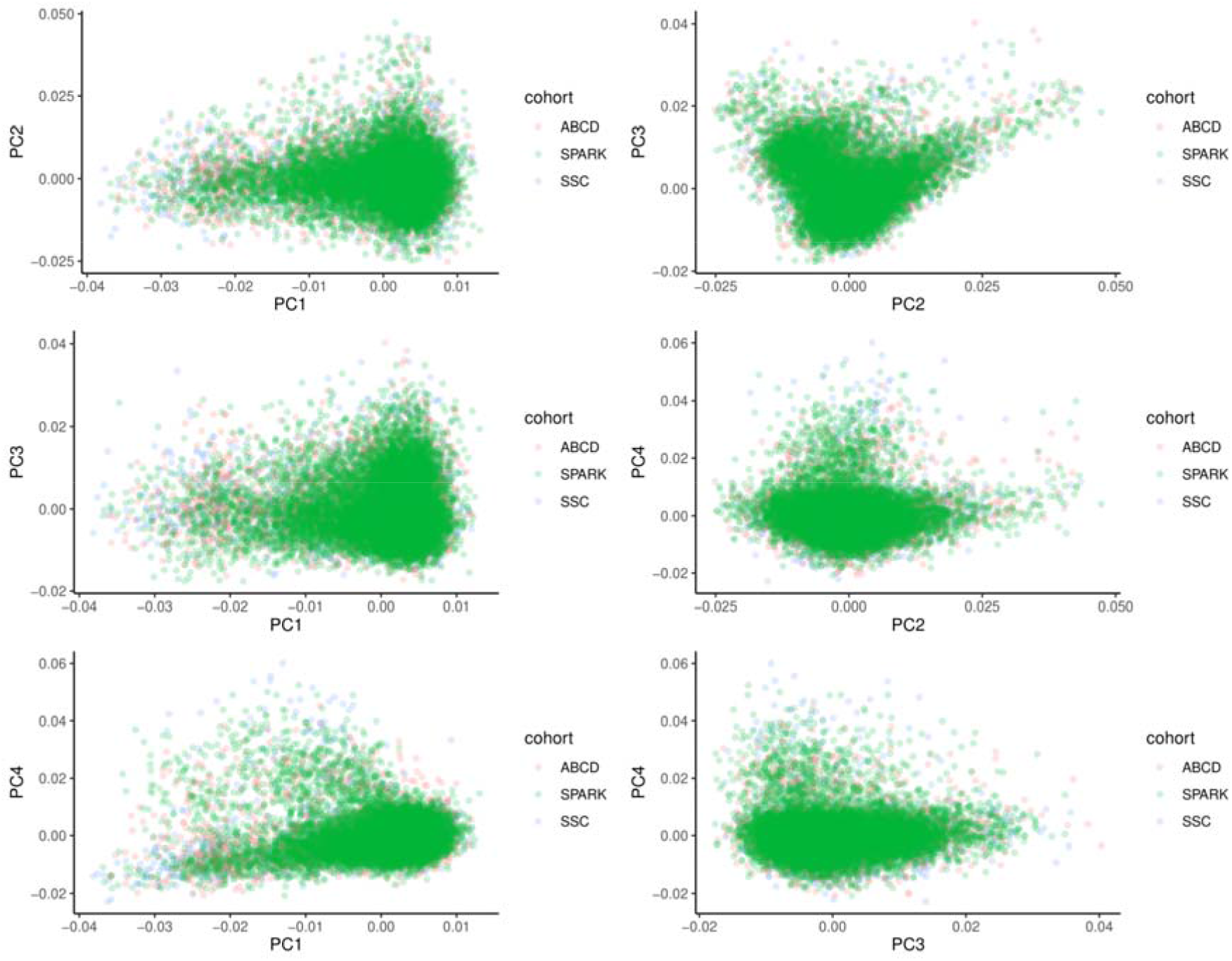
Distribution of individuals of European ancestries in SPARK, SSC, and ABCD by genetic principal components. Individuals of predominantly European ancestries from the SPARK, ABCD, and SSC cohorts plotted based on the first four genetic principal components. Visual inspection of the plots identified substantial alignment between the three cohorts in the principal component space.

## Footnotes

1# We use identity first language as this is preferred by the autistic community.Further information is available at: <u>how to talk about autism</u>

2# Throughout the manuscript, to orient the reader, we indicate associations where PGS are the independent variable with Beta_PGS_, and where high-impact de novo variants are the independent variable with Beta_denovo_

## References

1. Lai, M.-C., Lombardo, M. V. & Baron-Cohen, S. Autism. Lancet (2013) doi:10.1016/S0140-6736(13)61539-1.

2. American Psychiatric Association. Diagnostic and statistical manual of mental disorders (5th ed.). (2013).

3. Lord, C., et al. Autism spectrum disorder. Nat Rev Dis Primers 6, 5 (2020).

4. Geschwind, D. H. Advances in autism. Annu. Rev. Med. 60, 367–380 (2009).

5. Mandell, D. S., Novak, M. M. & Zubritsky, C. D. Factors associated with age of diagnosis among children with autism spectrum disorders. Pediatrics 116, 1480–1486 (2005).

6. Kanne, S. M. et al. The role of adaptive behavior in autism spectrum disorders: implications for functional outcome. J. Autism Dev. Disord. 41, 1007–1018 (2011).

7. Lai, M.-C. & Szatmari, P. Sex and gender impacts on the behavioural presentation and recognition of autism. Curr. Opin. Psychiatry 33, 117–123 (2020).

8. Warrier, V. et al. Elevated rates of autism, other neurodevelopmental and psychiatric diagnoses, and autistic traits in transgender and gender-diverse individuals. Nat. Commun. 11, 3959 (2020).

9. Frazier, T. W. et al. Demographic and clinical correlates of autism symptom domains and autism spectrum diagnosis. Autism 18, 571–582 (2014).

10. Havdahl, K. A. et al. Multidimensional Influences on Autism Symptom Measures: Implications for Use in Etiological Research. J. Am. Acad. Child Adolesc. Psychiatry 55, 1054–1063.e3 (2016).

11. Havdahl, A. et al. Genetic contributions to autism spectrum disorder. Psychol. Med. 1–14 (2021).

12. Warrier, V. et al. Social and non-social autism symptoms and trait domains are genetically dissociable. Commun Biol 2, 328 (2019).

13. Robinson, E. B., Lichtenstein, P., Anckarsäter, H., Happé, F. & Ronald, A. Examining and interpreting the female protective effect against autistic behavior. Proceedings of the National Academy of Sciences 110, 5258–5262 (2013).

14. Weiner, D. J. et al. Polygenic transmission disequilibrium confirms that common and rare variation act additively to create risk for autism spectrum disorders. Nat. Genet. (2017) doi:10.1038/ng.3863.

15. Robinson, E. B. et al. Genetic risk for autism spectrum disorders and neuropsychiatric variation in the general population. Nat. Genet. 48, 552–555 (2016).

16. Grove, J. et al. Identification of common genetic risk variants for autism spectrum disorder. Nat. Genet. 51, 431–444 (2019).

17. Satterstrom, F. K. et al. Large-Scale Exome Sequencing Study Implicates Both Developmental and Functional Changes in the Neurobiology of Autism. Cell 180, 568–584.e23 (2020).

18. Chaste, P. et al. A genome-wide association study of autism using the Simons Simplex Collection: Does reducing phenotypic heterogeneity in autism increase genetic homogeneity? Biol. Psychiatry 77, 775–784 (2015).

19. Antaki, D. et al. A phenotypic spectrum of autism is attributable to the combined effects of rare variants, polygenic risk and sex. medRxiv (2021).

20. Buja, A. et al. Damaging de novo mutations diminish motor skills in children on the autism spectrum. Proc. Natl. Acad. Sci. U. S. A. 115, E1859–E1866 (2018).

21. Bishop, S. L. et al. Identification of Developmental and Behavioral Markers Associated With Genetic Abnormalities in Autism Spectrum Disorder. Am. J. Psychiatry 174, 576–585 (2017).

22. Happé, F., Ronald, A. & Plomin, R. Time to give up on a single explanation for autism. Nat. Neurosci. 9, 1218–1220 (2006).

23. Frazier, T. W. et al. Validation of Proposed DSM-5 Criteria for Autism Spectrum Disorder. J. Am. Acad. Child Adolesc. Psychiatry 51, 28–40.e3 (2012).

24. Lai, M.-C., Lombardo, M. V., Auyeung, B., Chakrabarti, B. & Baron-Cohen, S. Sex/gender differences and autism: setting the scene for future research. J. Am. Acad. Child Adolesc. Psychiatry 54, 11–24 (2015).

25. Werling, D. M. & Geschwind, D. H. Sex differences in autism spectrum disorders. Curr. Opin. Neurol. 26, 146–153 (2013).

26. Kosmicki, J. A. et al. Refining the role of de novo protein-truncating variants in neurodevelopmental disorders by using population reference samples. Nat. Genet. 49, 504–510 (2017).

27. Kaplanis, J. et al. Evidence for 28 genetic disorders discovered by combining healthcare and research data. Nature 586, 757–762 (2020).

28. Lam, K. S. L. & Aman, M. G. The Repetitive Behavior Scale-Revised: Independent Validation in Individuals with Autism Spectrum Disorders. J. Autism Dev. Disord. 37, 855–866 (2007).

29. Rutter, M., Bailey, A. & Lord, C. SCQ: The Social Communication Questionnaire. (2003).

30. Fischbach, G. D. & Lord, C. The Simons Simplex Collection: A Resource for Identification of Autism Genetic Risk Factors. Neuron 68, 192–195 (2010).

31. SPARK Consortium. Electronic address, P., et al. SPARK: A US Cohort of 50,000 Families to Accelerate Autism Research. Neuron 97, 488–493 (2018).

32. Jones, R. M. et al. How interview questions are placed in time influences caregiver description of social communication symptoms on the ADI-R. Journal of Child Psychology and Psychiatry vol. 56 577–585 (2015).

33. Peça, J. et al. Shank3 mutant mice display autistic-like behaviours and striatal dysfunction. Nature 472, 437–442 (2011).

34. Katayama, Y. et al. CHD8 haploinsufficiency results in autistic-like phenotypes in mice. Nature 537, 675–679 (2016).

35. Hoffmann, T. J. et al. Evidence of reproductive stoppage in families with autism spectrum disorder: a large, population-based cohort study. JAMA Psychiatry 71, 943–951 (2014).

36. Lai, M.-C. & Baron-Cohen, S. Identifying the lost generation of adults with autism spectrum conditions. The Lancet Psychiatry 2, 1013–1027 (2015).

37. Clarke, T.-K. et al. Common polygenic risk for autism spectrum disorder (ASD) is associated with cognitive ability in the general population. Mol. Psychiatry 21, 419–425 (2015).

38. Myers, S. M. et al. Insufficient Evidence for ‘Autism-Specific’ Genes. Am. J. Hum. Genet. 106, 587–595 (2020).

39. Thormann, A. et al. Flexible and scalable diagnostic filtering of genomic variants using G2P with Ensembl VEP. Nat. Commun. 10, 2373 (2019).

40. Jacquemont, S. et al. A Higher Mutational Burden in Females Supports a ‘Female Protective Model’ in Neurodevelopmental Disorders. Am. J. Hum. Genet. 94, 415–425 (2014).

41. Sanders, S. J. et al. Insights into Autism Spectrum Disorder Genomic Architecture and Biology from 71 Risk Loci Article Insights into Autism Spectrum Disorder Genomic Architecture and Biology from 71 Risk Loci. Neuron 87, 1215–1233 (2015).

42. Study, D. D. D. & Deciphering Developmental Disorders Study. Prevalence and architecture of de novo mutations in developmental disorders. Nature vol. 542 433–438 (2017).

43. Wigdor, E. M. et al. The female protective effect against autism spectrum disorder. medRxiv (2021).

44. Pirastu, N. et al. Genetic analyses identify widespread sex-differential participation bias. Nature Genetics vol. 53 663–671 (2021).

45. Loomes, R., Hull, L. & Mandy, W. P. L. What Is the Male-to-Female Ratio in Autism Spectrum Disorder? A Systematic Review and Meta-Analysis. J. Am. Acad. Child Adolesc. Psychiatry 56, 466–474 (2017).

46. Yang, J., Lee, S. H., Goddard, M. E. & Visscher, P. M. GCTA: a tool for genome-wide complex trait analysis. Am. J. Hum. Genet. 88, 76–82 (2011).

47. Yang, J. et al. Common SNPs explain a large proportion of the heritability for human height. Nat. Genet. 42, 565–569 (2010).

48. Golan, D., Lander, E. S. & Rosset, S. Measuring missing heritability: inferring the contribution of common variants. Proc. Natl. Acad. Sci. U. S. A. 111, E5272–81 (2014).

49. Klei, L. L. et al. Common genetic variants, acting additively, are a major source of risk for autism. Mol. Autism 3, 9 (2012).

50. Gaugler, T. et al. Most genetic risk for autism resides with common variation. Nat. Genet. 46, 881–885 (2014).

51. Gao, Z. et al. Overlooked roles of DNA damage and maternal age in generating human germline mutations. Proc. Natl. Acad. Sci. U. S. A. 116, 9491–9500 (2019).

52. Kong, A. et al. Rate of de novo mutations and the importance of father’s age to disease risk. Nature 488, 471–475 (2012).

53. Niemi, M. E. K. et al. Common genetic variants contribute to risk of rare severe neurodevelopmental disorders. Nature 562, 268–271 (2018).

54. Trost, B. et al. Genome-wide detection of tandem DNA repeats that are expanded in autism. Nature 586, 80–86 (2020).

55. Mitra, I. et al. Patterns of de novo tandem repeat mutations and their role in autism. Nature 589, 246–250 (2021).

56. Dudbridge, F. Power and Predictive Accuracy of Polygenic Risk Scores. PLoS Genet. 9, e1003348 (2013).

57. Happé, F. & Frith, U. Annual Research Review: Looking back to look forward - changes in the concept of autism and implications for future research. J. Child Psychol. Psychiatry 61, 218–232 (2020).

58. Revelle, W. & Revelle, M. W. Package ‘psych’. The comprehensive R archive network 337, 338 (2015).

59. Bishop, S. L., Havdahl, K. A., Huerta, M. & Lord, C. Subdimensions of social-communication impairment in autism spectrum disorder. J. Child Psychol. Psychiatry 57, 909–916 (2016).

60. Zheng, S. et al. Extracting Latent Subdimensions of Social Communication: A Cross-Measure Factor Analysis. J. Am. Acad. Child Adolesc. Psychiatry 60, 768–782.e6 (2021).

61. Grove, R., Begeer, S., Scheeren, A. M., Weiland, R. F. & Hoekstra, R. A. Evaluating the latent structure of the non-social domain of autism in autistic adults. Mol. Autism 12, 22 (2021).

62. Richler, J., Bishop, S. L., Kleinke, J. R. & Lord, C. Restricted and repetitive behaviors in young children with autism spectrum disorders. J. Autism Dev. Disord. 37, 73–85 (2007).

63. Heise, D. R. & Bohrnstedt, G. W. Validity, Invalidity, and Reliability. Sociol. Methodol. 2, 104– 129 (1970).

64. Bentler, P. M. Alpha, Dimension-Free, and Model-Based Internal Consistency Reliability. Psychometrika 74, 137–143 (2009).

65. Reise, S. P., Moore, T. M. & Haviland, M. G. Bifactor models and rotations: exploring the extent to which multidimensional data yield univocal scale scores. J. Pers. Assess. 92, 544–559 (2010).

66. Rosseel, Y. Lavaan: An R package for structural equation modeling and more. Version 0.5--12 (BETA). J. Stat. Softw. 48, 1–36 (2012).

67. Geschwind, D. H. et al. The autism genetic resource exchange: a resource for the study of autism and related neuropsychiatric conditions. Am. J. Hum. Genet. 69, 463–466 (2001).

68. Charman, T. et al. The EU-AIMS Longitudinal European Autism Project (LEAP): clinical characterisation. Mol. Autism 8, 27 (2017).

69. Gibbs, R. A. et al. A global reference for human genetic variation. Nature 526, 68–74 (2015).

70. McInnes, L., Healy, J., Saul, N. & Großberger, L. UMAP: Uniform Manifold Approximation and Projection. Journal of Open Source Software vol. 3 861 (2018).

71. Conomos, M. P. & Thornton, T. GENetic EStimation and inference in structured samples (GENESIS): statistical methods for analyzing genetic data from samples with population structure and/or relatedness. R package version 2, (2016).

72. Manichaikul, A. et al. Robust relationship inference in genome-wide association studies. Bioinformatics 26, 2867–2873 (2010).

73. Howie, B. N., Fuchsberger, C., Stephens, M., Marchini, J. & Abecasis, G. R. Fast and accurate genotype imputation in genome-wide association studies through pre-phasing. Nat. Genet. 44, 955–959 (2012).

74. McCarthy, S. et al. A reference panel of 64,976 haplotypes for genotype imputation. Nat. Genet. 48, 1279–1283 (2016).

75. Taliun, D. et al. Sequencing of 53,831 diverse genomes from the NHLBI TOPMed Program. Nature 590, 290–299 (2021).

76. Warrier, V. et al. Gene–environment correlations and causal effects of childhood maltreatment on physical and mental health: a genetically informed approach. The Lancet Psychiatry 8, 373– 386 (2021).

77. Lee, J. J. et al. Gene discovery and polygenic prediction from a genome-wide association study of educational attainment in 1.1 million individuals. Nat. Genet. 50, 1112–1121 (2018).

78. Savage, J. E. et al. Genome-wide association meta-analysis in 269,867 individuals identifies new genetic and functional links to intelligence. Nat. Genet. 1 (2018).

79. Ripke, S., Walters, J. T. R., O’Donovan, M. C., the Psychiatric Genomics Consortium, S. W. G. of & Others. Mapping genomic loci prioritises genes and implicates synaptic biology in schizophrenia. MedRxiv (2020).

80. Ge, T., Chen, C.-Y., Ni, Y., Feng, Y.-C. A. & Smoller, J. W. Polygenic prediction via Bayesian regression and continuous shrinkage priors. Nat. Commun. 10, 1776 (2019).

81. Pain, O. et al. Evaluation of polygenic prediction methodology within a reference-standardized framework. PLoS Genet. 17, e1009021 (2021).

82. Samocha, K. E., Kosmicki, J. A. & Karczewski, K. J. Regional missense constraint improves variant deleteriousness prediction. BioRxiv (2017).

83. Karczewski, K. J. et al. Author Correction: The mutational constraint spectrum quantified from variation in 141,456 humans. Nature 590, E53 (2021).

84. Lord, C. et al. Autism diagnostic observation schedule: a standardized observation of communicative and social behavior. J. Autism Dev. Disord. 19, 185–212 (1989).

85. Lord, C. et al. Autism Diagnostic Interview-Revised: A revised version of a diagnostic interview for caregivers of individuals with possible pervasive developmental disorders. J. Autism Dev. Disord. 24, 659–685 (1994).

86. Constantino, J. N. & Gruber, C. P. *Social responsiveness scale: SRS-2*. (Western Psychological Services Torrance, CA, 2012).

87. Sparrow, S. S., Balla, D. A., Cicchetti, D. V. & Harrison, P. L. Vineland adaptive behavior scales. (1984).

88. Wilson, B. N., Kaplan, B. J., Crawford, S. G. & Roberts, G. The developmental coordination disorder questionnaire 2007 (DCDQ’07). Administrative manual for the DCDQ107 with psychometric properties 267–272 (2007).

89. Ripley, B. et al. Package ‘mass’. Cran r 538, 113–120 (2013).

90. Bates, D., Sarkar, D., Bates, M. D. & Matrix, L. The lme4 package. R package version 2, 74 (2007).

91. Diedenhofen, B. & Musch, J. cocor: a comprehensive solution for the statistical comparison of correlations. PLoS One 10, e0121945 (2015).

92. Peyrot, W. J., Boomsma, D. I., Penninx, B. W. J. H. & Wray, N. R. Disease and polygenic architecture: Avoid trio design and appropriately account for unscreened control subjects for common disease. Am. J. Hum. Genet. 98, 382–391 (2016).

93. Baron-Cohen, S. The hyper-systemizing, assortative mating theory of autism. Prog. Neuropsychopharmacol. Biol. Psychiatry 30, 865–872 (2006).

94. Lee, S. H., Wray, N. R., Goddard, M. E. & Visscher, P. M. Estimating missing heritability for disease from genome-wide association studies. Am. J. Hum. Genet. 88, 294–305 (2011).

95. Maenner, M. J. et al. Prevalence of Autism spectrum disorder among children aged 8 years - Autism and Developmental Disabilities Monitoring Network, 11 sites, United States, 2016. MMWR Surveill. Summ. 69, 1–12 (2020).

96. Satterstrom, F. K. et al. Autism spectrum disorder and attention deficit hyperactivity disorder have a similar burden of rare protein-truncating variants. Nat. Neurosci. 22, 1961–1965 (2019).

